# Innate immune activation restricts priming and protective efficacy of the radiation-attenuated PfSPZ malaria vaccine

**DOI:** 10.1101/2021.10.08.21264577

**Authors:** Leetah Senkpeil, Jyoti Bhardwaj, Morgan Little, Prasida Holla, Aditi Upadhye, Phillip A. Swanson, Ryan E. Wiegand, Michael D. Macklin, Kevin Bi, Barbara J. Flynn, Ayako Yamamoto, Erik L. Gaskin, D. Noah Sather, Adrian L. Oblak, Edward Simpson, Hongyu Gao, W. Nicholas Haining, Kathleen B. Yates, Xiaowen Liu, Kephas Otieno, Simon Kariuki, Xiaoling Xuei, Yunlong Liu, Rafael Polidoro, Stephen L. Hoffman, Martina Oneko, Laura C. Steinhardt, Nathan W. Schmidt, Robert A. Seder, Tuan M. Tran

## Abstract

Baseline innate immune signatures can influence protective immunity following vaccination. Here, we used systems transcriptional analysis to assess the molecular mechanisms underlying differential immunogenicity and protective efficacy results of a clinical trial of the radiation-attenuated whole sporozoite PfSPZ Vaccine in African infants. Innate immune activation and myeloid signatures at pre-vaccination baseline correlated with protection from *Plasmodium falciparum* infection in placebo controls, while the same signatures predicted susceptibility to infection among infants who received the highest and most protective dose of the PfSPZ Vaccine. Machine learning identified monocytes and an antigen presentation signature as pre-vaccination features predictive of malaria infection after highest-dose PfSPZ vaccination. Consistent with these human data, innate stimulation *in vivo* conferred protection against malaria infection in mice while diminishing the CD8^+^ T cell response to radiation-attenuated sporozoites. These data establish a dichotomous role of innate stimulation for malaria protection and induction of protective immunity of whole-sporozoite malaria vaccines.

## INTRODUCTION

Malaria, caused by mosquito-borne *Plasmodium* parasites, was responsible for 409,000 deaths in 2019, primarily due to *Plasmodium falciparum* (Pf) in sub-Saharan African children (World Health Organization, 2020). Effective and broad implementation of malaria control measures has greatly reduced global malaria morbidity and mortality over the last two decades, However, progress has stalled since 2015, underscoring the need for a highly effective vaccine for malaria prevention (World Health Organization, 2020). Natural immunity that prevents Pf infection at the pre-erythrocytic stage is not reliably acquired even after years of parasite exposure (Tran et al., 2013). “Pre-erythrocytic” malaria vaccines that target the *Plasmodium* parasite prior to the symptomatic blood-stage, aim to induce sterilizing immunity that inhibits sporozoite entry into hepatocytes or impedes liver-stage development. The most advanced malaria vaccine to date, RTS,S, is a subunit vaccine comprised of an immunodominant B-cell epitope (the NANP repeat sequence) and T-cell epitopes from the *P. falciparum* circumsporozoite protein (CSP) fused to hepatis B surface antigen and formulated with the adjuvant AS01. In a Phase 3 clinical trial of 8,922 5-17-month-old infants in sub-Saharan Africa, a four-dose regimen of RTS,S/AS01 conferred 36.3% vaccine efficacy (VE) against clinical malaria over four years (Rts, 2015). Recently, a next-generation RTS,S-like vaccine, R21, demonstrated 77% VE against clinical malaria at 6 months, the time of the most intense malaria transmission in a Phase 2b trial of 450 infants aged 5-17 months (Datoo et al., 2021).

Immunization with radiation-attenuated sporozoites (RAS) represents another approach for inducing sterile protection against *Plasmodium* infection. Initially demonstrated in a rodent malaria model (Nussenzweig et al., 1967), RAS immunization was later shown to be protective in humans when delivered by mosquito bite (Clyde, 1975). Induction of highly effective sterilizing immunity by this approach requires that RAS undergo arrested intra-hepatocytic development after initial infection of liver cells (Nganou-Makamdop and Sauerwein, 2013). Aseptic, purified, live, non-replicating, radiation-attenuated cryopreserved Pf sporozoites (referred henceforth as PfSPZ Vaccine) developed for direct inoculation in humans showed ∼60-100% VE against Pf parasitemia up to 14 months after challenge by controlled human malaria infection (CHMI) with homologous parasites in malaria-naïve North American volunteers (Seder et al., 2013). When delivered via direct venous inoculation (DVI), the PfSPZ Vaccine confers malaria protection by inducing circulating CSP-specific Abs and both peripheral and liver-resident Pf-specific T-cell responses (Ishizuka et al., 2016; Lyke et al., 2017; Seder et al., 2013), with the latter being critical for durable sterilizing immunity (Epstein et al., 2011; Weiss et al., 1988). Evidence suggests that γδ (Vγ9Vδ2) T cells may be important for mediating priming of protective CD4^+^ and CD8^+^ T-cell responses during PfSPZ vaccination. The frequency of Vγ9Vδ2 T cells prior to immunization with PfSPZ Vaccine has been shown to correlate with the induction of Pf-specific T cells and protective outcomes and expand with PfSPZ immunization (Ishizuka et al., 2016; Zaidi et al., 2017).

A PfSPZ Vaccine field trial conducted in malaria-exposed Malian adults demonstrated a VE of 28% against microscopy-detectable Pf parasitemia by proportional analysis during six months of intense, natural Pf transmission (Sissoko et al., 2017). Identical PfSPZ Vaccine regimens were significantly less immunogenic in terms of both Pf-specific Ab and CD4^+^ T cell responses in malaria-exposed African adults when compared to malaria-naïve adults (Ishizuka et al., 2016; Jongo et al., 2019; Sissoko et al., 2017), suggesting that long-term exposure to malaria may limit both vaccine immunogenicity and efficacy. Infants and young children in sub-Sahara Africa are relatively malaria-inexperienced but also represent the age groups at the greatest risk for severe malaria and death. Based on the hypothesis that less prior malaria exposure may enhance vaccine responsiveness, a phase 2 double-blinded, randomized, placebo-controlled trial of the PfSPZ Vaccine was conducted in Kenyan infants from January 2017 to August 2018 (Oneko et al., 2021). Groups of 84 infants received 4.5×10^5^, 9.0×10^5^, 1.8×10□ PfSPZ Vaccine or normal saline (placebo) three times at 8-week intervals (**Figure 1A**). Although there was no significant VE against Pf parasitemia for any dose group at the primary endpoint of 6 months, VE in the highest dose group was 41.1% at 3 months after the last dose. PfSPZ vaccination generated robust CSP-specific Abs that correlated with protection but did not generate detectable Pf-specific T-cell responses in any dose group. The similar outcomes across treatment groups provided an opportunity to investigate the molecular differences between infants who did and did not effectively respond to the PfSPZ Vaccine as measured by immunogenicity or by protection against incident Pf parasitemia during the surveillance period. To gain better insight into the molecular mechanisms underlying the immunogenicity and efficacy results from this pediatric vaccine trial, we used pre- and post-immunization blood samples from infants to conduct a systems analysis that integrated whole-blood transcriptomic profiling with Pf-specific Ab, immunophenotyping, plasma cytokine, and clinical data (**Figure 1A**; **Figure S1**).

**Figure 1.**
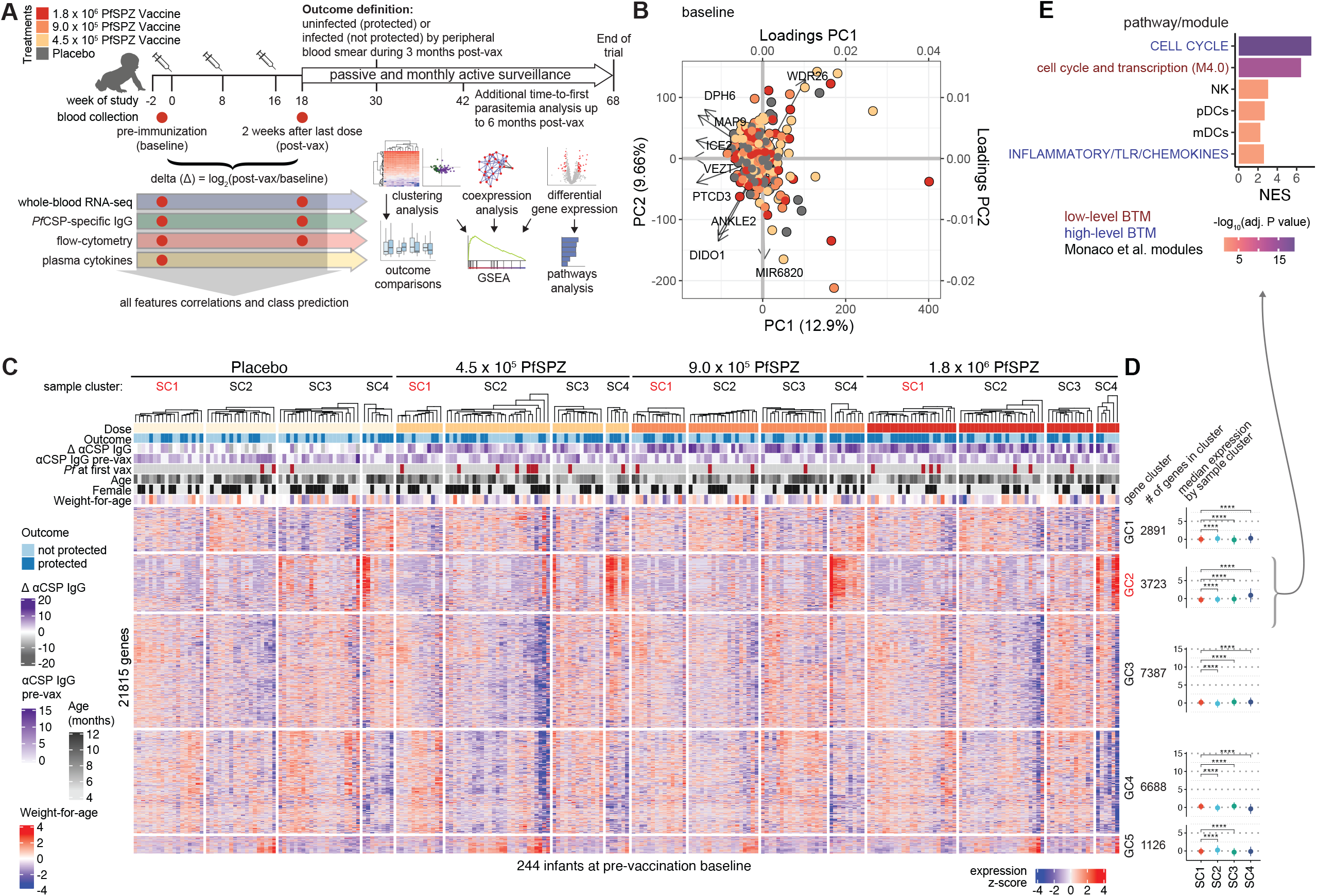
Variation in baseline transcriptomes defined by gene modules. (A) Immunization, blood collection, and surveillance schedule and general experimental and analysis workflow. (B) Biplot with top principal components (PC) and associated loadings for all transcriptomes at baseline (n=244). (C) Hierarchical clustering heatmap of baseline transcriptomes grouped by treatment with column annotations for outcome, CSP-specific IgG responses, and subject characteristics. Partitioning around medoids and Euclidean distance metric were used for clustering with k=4 and k=5 for sample clusters (SC) and gene clusters (GC), respectively. (D) Comparison of median expression of genes within each gene cluster between the sample clusters. (E) Gene set enrichment analysis of genes in GC2 ranked by median expression across all samples (minimum gene set size=20, FDR<5%). NES = normalized enrichment score.

## RESULTS

### Clinical outcomes and overview of participants and samples

In the clinical trial, the primary outcome was presence (not protected; NP) or absence (protected; P) of microscopy-detectable Pf infection through 6 months of surveillance after the third dose of vaccine or placebo (Oneko et al., 2021). However, for the current study, we used the secondary outcomes of protection through 3 months of surveillance or days to first Pf parasitemia up to 168 days (∼6 months) (**Figure 1A**). Of the 336 participants in the parent KSPZV1 trial, 258 had whole-blood RNA of sufficient quality for analysis from at least one time point (**Figure S1A**). Characteristics between NP and P infants by treatment were similar except for differences in malaria transmission at the two recruitment sites (**Figure S1B**). Serum, plasma and PBMCs used for subsequent immune profiling assays were evenly distributed across treatment groups (**Figure S1C**).

### Variation in baseline transcriptomes defined by gene modules

To assess the overall variation in gene expression profiles at baseline across all treatment groups and outcomes pre-immunization, we performed clustering analysis of baseline blood transcriptomes from 244 of the 336 infants enrolled in the vaccine trial (**Figure 1A**; **Figure S1A**). Principal component (PC) analysis did not demonstrate distinct clustering by vaccine dose group; however, the top genes driving variation along PC1 and PC2 include *DPH6*, a gene involved in the regulation of several innate and adaptive cell lineages (Abeler-Dorner et al., 2020); *ICE2*/*NARG2*, a gene associated with asthma risk (Schoettler et al., 2019); and *WDR26*, an erythroid enriched gene that regulates nuclear condensation during erythropoeisis (Zhen et al., 2020) (**Figure 1B**). Unsupervised clustering of pre-immunization blood transcriptomes revealed a sample cluster (SC1) within the 1.8×10^6^ PfSPZ group that was significantly overrepresented by P infants (**Figure 1C; Figure S2A-B**). Among SC1 infants receiving PfSPZ Vaccine, neither pre- nor post-immunization CSP-specific IgG differed between the outcome groups (**Figure S2C-D**). In contrast, P infants in SC2-4 demonstrated greater CSP-specific IgG responses to either 4.5×10^5^ PfSPZ or 1.8×10^6^ PfSPZ (**Figure S2D**). Among SC1 infants who received placebo, P infants (who never had microscopy-detectable parasitemia during 3 months of surveillance) began the study with lower CSP-specific IgG but demonstrated greater IgG responses to CSP during the vaccination period when compared to NP infants who became parasitemic (**Figure S2C-D**). Weight-for-age z-score was significantly decreased in P infants relative to NP infants only within SC1 of the 4.5×10^5^ PfSPZ group (**Figure S2E**). No other significant differences were observed between the outcome groups within SC1 for any treatment group for CSP-specific IgG, presence of Pf parasitemia at the first vaccination, age, gender, or nutritional status (**Figure S2C-H**). Across all treatment groups, gene expression within gene clusters (GC) for infants in SC1 significantly differed from expression in the other sample clusters, with expression of genes in GC2 being significantly decreased relative to SC2, SC3, or SC4 (**Figure 1D**). Enrichment analysis using blood-transcription modules (BTMs) (Li et al., 2014) and modules derived from a flow-sorted RNA-seq dataset (Monaco et al., 2019) revealed that genes within GC2 are related to cell cycle, natural killer (NK) cells, dendritic cells (DCs), and inflammation/toll-like receptors (TLRs)/chemokines (**Figure 1E**). Taken together, these broad assessments of variation across the baseline blood transcriptomes suggest decreased innate inflammation prior to vaccination may define protective responses to high-dose PfSPZ Vaccine in a manner that is independent of the Ab response to sporozoites.

### Innate activation, myeloid, and erythroid signatures at baseline distinguish protective outcomes

To determine the relationship between genes with highly correlated expression at baseline and protective outcomes irrespective of vaccination, we constructed data-driven modules from the 244 pre-immunization transcriptomes using weighted gene co-expression network analysis (WGCNA) and correlated them to parasitemia and CSP-specific IgG variables. Three mutually exclusive modules driven by the hub genes *RIOK3, CSDE1*, and *SEC62* negatively correlated with the protected (uninfected) outcome and positively correlated with both pre-immunization anti-CSP Abs and presence of Pf parasitemia at the first vaccination (**Figure 2A**). RIOK3 has been shown to attenuate the innate immune response by mediating the phosphorylation of the RNA-sensor MDA5 (Takashima et al., 2015). CSDE1 is strongly upregulated in erythropoiesis and its loss inhibits erythroid proliferation and differentiation (Horos et al., 2012). *SEC62* encodes a translocon component that is critical for maintaining ER homeostasis during stress recovery (Fumagalli et al., 2016). A single module positively correlated with protection and was led by the hub-gene *EFHD2*, which encodes a calcium-binding protein that was shown to positively regulate the B-cell-receptor induced intracellular calcium response (Kroczek et al., 2010) and is required for human T-cell mediated cytotoxicity (Peled et al., 2018) (**Figure 2A**). Network graphs of nodal correlations revealed that modules negatively correlated with protection were tightly linked to each other and only weakly linked to the protection-associated *EFHD2* module (**Figure 2B**). To gain further insight into these networks, GSEA was applied to genes within each WGCNA-derived module. Although none were enriched in genes within BTMs or Monaco gene sets, using BloodGen3Module (Rinchai et al., 2021), the hallmark collection (Liberzon et al., 2015), and KEGG pathways (Kanehisa et al., 2021), the *EFHD2*-hubbed module was enriched for genes related to chemokine signaling, regulation of actin cytoskeleton, Fcγ receptor-mediated phagocytosis, and inflammation. In contrast, two modules that negatively correlated with protection were enriched in genes related to heme metabolism and erythrocytes (**Figure 2C**).

**Figure 2.**
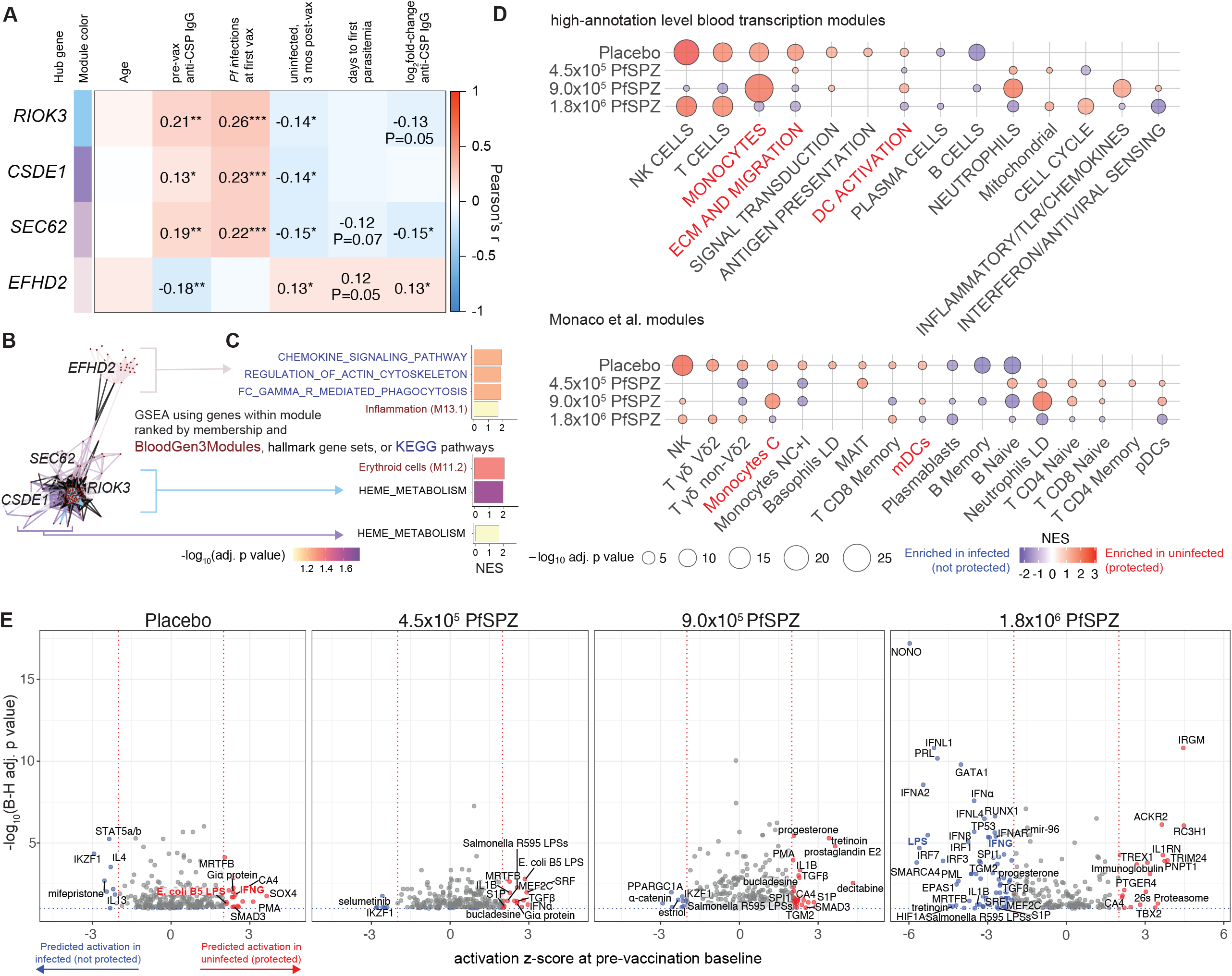
Innate activation, myeloid, and erythroid signatures at baseline distinguish protective outcomes. (A) Correlations between module eigengenes, obtained by weighted gene correlation network analysis, with age, baseline factors, and outcomes. Only modules that significantly correlated with the uninfected outcome (P<0.05) are shown. (B) Network graphs of significant modules containing nodes (red dots) and edges (lines) meeting minimum thresholds (**see Methods**). Correlations between nodes in different modules are shown as black edges. (C) GSEA of genes within modules that significant correlated with outcome using blood transcription modules, BloodGen3Modules, and KEGG pathways as gene sets (BH-adjusted p<0.10). The *SEC62*-hubbed module did not show significant enrichment for any gene set. (D) GSEA of genes ranked by differential expression between uninfected (protected) and infected (not protected) infants for each PfSPZ Vaccine dose group. Only modules with a B-H-adjusted p<0.20 are shown. Modules in which enrichment is reversed between placebo and 1.8×10^6^ PfSPZ Vaccine groups are in red text. (E) Volcano plots of upstream regulators predicted to be differentially activated between P and NP infants (|z-score|>2 and B-H-adjusted p<0.05) for each treatment. Predicted upstream regulators that are reversed between the placebo and 1.8×10^6^ PfSPZ Vaccine groups are labeled in color. B-H = Benjamini-Hochberg.

We next directly compared baseline transcriptomic profiles between the 3-month outcomes for each of the dose groups using differential gene expression (DGE) analysis. Although no differentially expressed genes were observed between outcomes for any of the dose groups at a false discovery rate (FDR)<20% (**Table S1**), GSEA revealed significant differences between P and NP infants. Cytotoxic lymphocyte signatures inclusive of NK cells, γδ T cells, and CD8 T cells were positively enriched in P relative to NP in both placebo and 1.8×10^6^ PfSPZ dose group, a finding not consistently observed for the other PfSPZ dose groups (**Figures 2D and S3**). Myeloid signatures, inclusive of monocytes and DCs (M81, S4, M11.0, M165), and innate inflammation signatures (“TNF via NFκB signaling”, “IL6-JAK-STAT3 signaling”, “interferon (IFN) γ response”) were positively enriched in P relative to NP in the placebo group (**Figures 2D and S3**). Notably, however, the same myeloid and innate inflammation signatures were negatively enriched in P relative to NP in the 1.8×10^6^ PfSPZ group, suggesting an interaction between the presence of these signatures at baseline and natural (placebo) versus vaccine-induced protective outcome. Other signatures that were differentially enriched in a reciprocal manner between placebo and 1.8×10^6^ PfSPZ include “cell adhesion” (M51), “myogenesis”, “epithelial mesenchymal transition”, “hedgehog signaling”, “prostanoids” (M8.2), and “platelet/prostaglandins” (M16.64) (**Figure S3A-C**). Interestingly, erythroid modules were consistently enriched in NP at baseline across all four treatment groups (**Figure S3C**), suggesting that baseline erythropoiesis predicted future susceptibility to Pf parasitemia regardless of vaccination regimen. The reciprocal differential enrichment observed between the placebo and 1.8×10^6^ PfSPZ groups was corroborated by upstream regulator analysis, which predicts regulators based on differential expression of downstream targets (Kramer et al., 2014) (**Figure 2E**). The innate inflammatory regulators lipopolysaccharide (LPS) and IFNG were predicted to be activated at baseline in P relative to NP for placebo, 4.5×10^5^ PfSPZ, and 9.0×10^5^ PfSPZ. In contrast, LPS and IFNG, along with type I, type III interferons and interferon regulatory factors (IRFs), were predicted to be activated in NP relative to P for the 1.8×10^6^ PfSPZ group, with greater absolute z-scores and significance than observed in the other dose groups (**Figure 2E**). Notable regulators also predicted to be activated in NP within the 1.8×10^6^ PfSPZ group include NONO, a nuclear protein which can promote cGAS-mediated innate immune activation in DCs and macrophages (Lahaye et al., 2018), the erythroid transcription factor GATA1, and the inflammatory cytokine IL-1β. To validate these findings, we analyzed whole-blood RNA-seq data collected from malaria-naïve adult volunteers from the VRC312 and VRC314 PfSPZ Vaccine trials (Ishizuka et al., 2016; Seder et al., 2013), limiting to regimens with uniform PfSPZ Vaccine doses administered by DVI (**Figure S4A; Table S1**). In these studies, protection was defined as absence of Pf parasitemia during 4 weeks of surveillance post-CHMI challenge. Consistent with the Kenyan infants who received 1.8×10^6^ PfSPZ, myeloid (monocytes, DCs) and innate inflammation signatures (interferon, inflammatory/TLR/chemokines) were enriched prior to vaccination in individuals who would later develop parasitemia after challenge (**Figure S4B**). Moreover, LPS and inflammatory cytokines (IL-1β, IL-6, and TNF) were predicted to be activated in VRC312 but not VRC314 (**Figure S4C**). These data suggest that innate immune activation of monocytes and dendritic cells prior to the initial dose impairs the efficacy of high-dose PfSPZ Vaccine but may be protective against natural infections in individuals receiving placebo or suboptimal PfSPZ Vaccine doses.

### Baseline monocyte and innate inflammatory signatures correlate with post-immunization CSP-specific B cell responses

To identify molecular signatures predictive of protective immune cell phenotypes, we first correlated time-to-first parasitemia up to 6 months post-immunization with cell subset frequencies at pre-immunization baseline or 2-week post-immunization as well as with the fold-difference between the two time points. These flow cytometry data were originally generated as part of the clinical study (Oneko et al., 2021). The immune phenotypes showing the strongest positive correlation with time-to-first parasitemia were post-immunization CSP-specific B cells (particularly CSP-specific memory B cells) and pre-immunization Vδ2 γδ T cells (particularly Vγ9^+^Vδ2^+^ γδ T cells), consistent with their protective associations observed in prior studies (Ishizuka et al., 2016; Jagannathan et al., 2017; Zaidi et al., 2017). We used post-immunization CSP-specific memory B cells as a surrogate for an effective adaptive response to the vaccine given their highly significant correlation with time-to-first parasitemia (**Figures 3A-B**). Pre-immunization genes that correlated with post-immunization CSP-specific memory B cells were positively enriched for signatures related to monocytes, plasmacytoid DCs, neutrophils, NK cells, and inflammation but negatively enriched for B-cell and interferon signatures (**Figure 3C**). We next looked at CSP-specific IgG reactivity, which was associated with protection in the vaccine trial (Oneko et al., 2021). In the 1.8×10^6^ PfSPZ group, CSP-specific IgG was significantly higher at baseline in NP versus P infants, suggesting that pre-existing anti-sporozoite Abs may inhibit the efficacy of the high-dose PfSPZ Vaccine (**Figure 3D**). When comparing fold-change of CSP-specific IgG responses to account for the variation in pre-existing Abs, higher vaccine-induced CSP-specific IgG responses were observed in P versus NP for both 4.5×10^5^ and 1.8×10^6^ PfSPZ groups, with the latter approaching significance (**Figure 3D**). Taking advantage of the bimodal CSP-specific IgG response to high-dose PfSPZ Vaccine, we performed DGE followed by enrichment analysis between high and low CSP-specific IgG responders within the 1.8×10^6^ PfSPZ group (**Table S2**; **Figures 3D-E**). High-CSP IgG responders were positively enriched for genes related to innate myeloid cells (monocytes, myeloid DCs, neutrophils), inflammatory/TLR/chemokines, antigen presentation, and, to a lesser extent, IFN/antiviral sensing prior to vaccination. In contrast, low-CSP IgG responders were positively enriched for lymphoid signatures, particularly cytotoxic signatures (NK cells, CD8^+^ memory T cells, non-Vδ2 γδ T cells) (**Figure 3E**). Taken together, these data suggest that innate immune activation in monocytes, DCs, and neutrophils at baseline enhances the vaccine-induced PfSPZ-specific Ab response.

**Figure 3.**
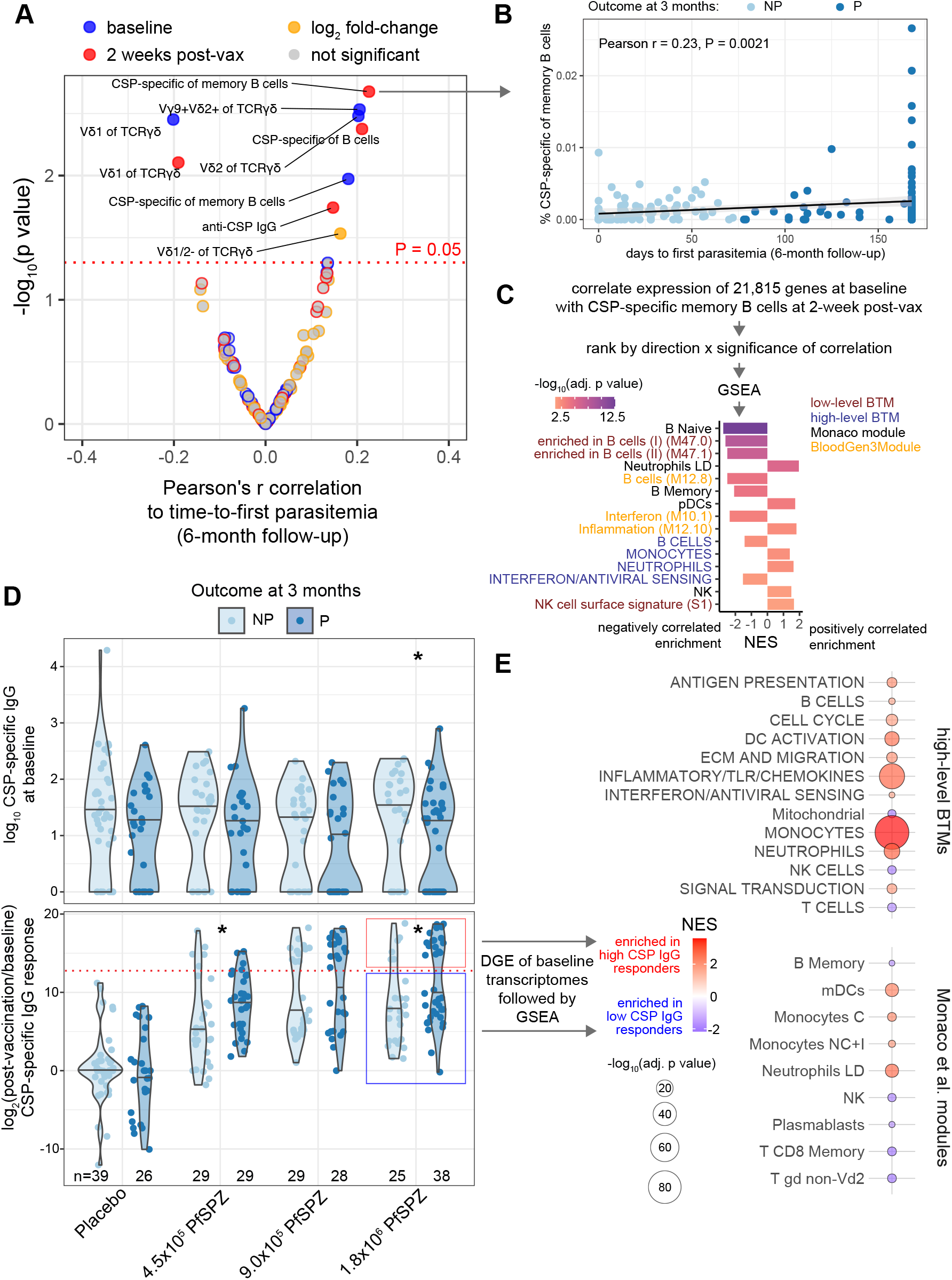
Baseline monocyte and innate inflammatory signatures correlate with post-immunization CSP-specific B cell responses. (A) Volcano plot of CSP-specific IgG and flow cytometry features at each timepoint or calculated as fold-change post-vaccination over baseline. (B) Correlation between CSP-specific memory B cells and time-to-first parasitemia up to 6 months (n=183) by presence (not protected, NP) or absence (protected, P) of parasitemia through 3 months of follow up. (C) GSEA using genes ranked by direction and significance of correlation between baseline expression and % CSP-specific of memory B cells at 2-weeks post-vaccination. (D) Violin plots showing CSP-specific IgG at baseline and as fold-change (2-weeks post-vaccination/baseline) by treatment and outcome at 3 months. *p<0.05 between outcomes within a treatment by Wilcoxon test. Red dotted line indicates threshold for high- and low-CSP IgG. (E) GSEA using genes ranked by direction and significance of differential gene expression (DGE) at baseline between infants immunized with 1.8×10^6^ PfSPZ who subsequently developed either a high or low CSP IgG response post-vaccination as defined in D. For C and E, only modules with a Benjamini-Hochberg-adjusted p<0.05 are shown. NES = normalized enrichment score.

### Peripheral gene signatures induced by high-dose PfSPZ vaccination predict protection from parasitemia

To assess the variation in blood transcription profiles during the vaccination period while accounting for inter-subject baseline differences, we examined paired changes in global gene expression at 2 weeks post-immunization relative to the pre-immunization baseline. Among the primary drivers of variation in transcriptional changes during the vaccination period were *PRKCD*, which encodes a signaling kinase that plays a central role in B-cell homeostasis and tolerance (Mecklenbrauker et al., 2002; Miyamoto et al., 2002); *STAT3*, which encodes for a IL-6/JAK-activated transcriptional regulator that plays a critical role in innate and adaptive immune responses (Johnson et al., 2018); *TFE3*, which is critical for T cell–dependent Ab responses (Huan et al., 2006); and *IL17RA*, which encodes the receptor for the proinflammatory cytokine IL-17 (**Figure 4A**). *ICE2*, which drove baseline transcriptomic variation (**Figure 1B**), also drove variation during the vaccination period along PC1 (**Figure 4A**). Unsupervised clustering of changes in blood transcriptomes during the vaccination period revealed a sample cluster (SC2) within the 1.8×10^6^ PfSPZ group that was overrepresented by P infants, a result that approached significance (**Figure 4B; Figure S5A**). In contrast, within the placebo group, P infants were proportionally underrepresented in SC2. Among SC2 infants in the 1.8×10^6^ PfSPZ group, CSP-specific IgG at baseline or fold-change post-immunization were similar between outcomes and not statistically different from the other sample clusters (**Figure S5A**). Overrepresentation of P infants within 1.8×10^6^ PfSPZ SC2 also could not be explained by differences in Pf infections at first vaccination or during the vaccination period (**Figure S5A**). Unsupervised clustering also revealed a highly variable cluster of genes (GC4) induced among infants in SC2 (**Figure 4B-C**). Highly variable genes within GC4 were related to integrin cell surface; collagen, TGFB family, extracellular matrix and migration, signal transduction, and inflammatory/TLR/chemokines (**Figure 4D**). The global analysis suggests that transcriptomic variation during the vaccination period may be explained by changes in genes related to cellular migration, signaling pathways, and both innate inflammatory and adaptive responses and that induction of these genes may influence the outcome based on the vaccination received.

**Figure 4.**
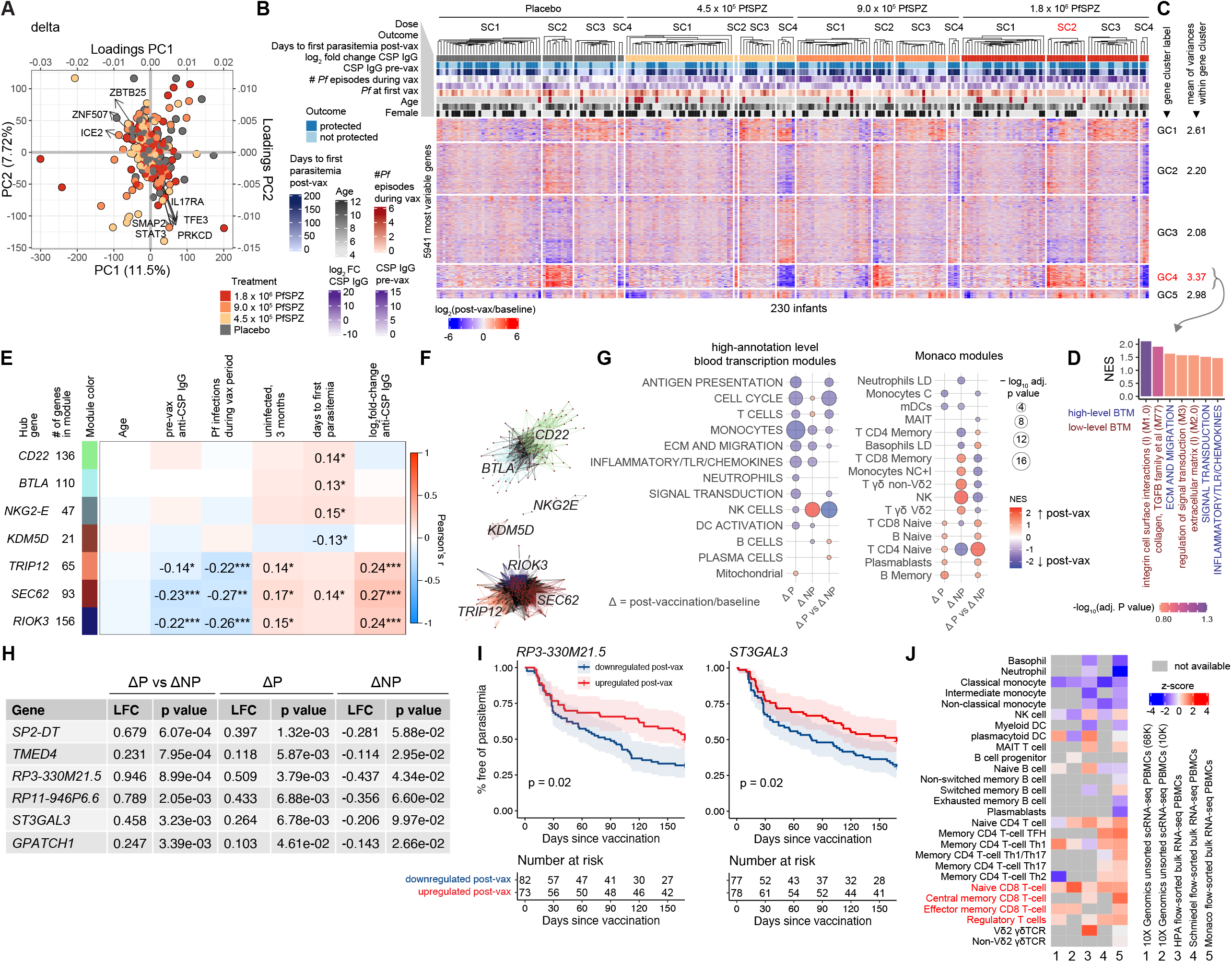
Peripheral gene signatures induced by high-dose PfSPZ vaccination predict protection from parasitemia. (A) Biplot showing the top principal components (PC) and associated loadings for “delta” transcriptomes determine as log_2_(post-vaccination/baseline) for 230 infants with paired data. (B) Hierarchical clustering heatmap of longitudinal transcriptomic changes for the top 25% most variable genes grouped by treatment with column annotations for outcome, CSP-specific IgG responses, and subject characteristics. Ward.D2 and Euclidean distance metric used for clustering samples (SC) and genes (GC). (C) Comparison of mean of variances within each gene cluster. (D) GSEA of genes within the most variable cluster GC4 ranked by mean variance across all samples (minimum gene set size = 5, only modules with BH-adjusted p<0.20 shown). (E) Correlations of the module eigengenes, obtained by weighted gene correlation network analysis of changes in gene expression (post-immunization/baseline) for all infants, with indicated variables. Only modules that significantly correlated with outcomes (p<0.05) are shown. (F) Network graphs of modules in E containing nodes (genes) and edges (correlations) with minimum thresholds. (G) GSEA of genes ranked by differential expression between post-vaccination vs. baseline (δ) within the protected (ΔP) or not protected (ΔNP) groups or between outcome groups adjusting for baseline (ΔP vs ΔNP) for infants receiving 1.8×10^6^ PfSPZ Vaccine. (H) Genes differentially expressed between ΔP and ΔNP (p<0.005) and within δP (p<0.05) in 1.8×10^6^ PfSPZ. (I) Kaplan-Meier plot of risk of parasitemia during 6 months of surveillance for PfSPZ-vaccinated infants with or without upregulation of indicated gene 2 weeks post-vaccination. Significance determined by log-rank analysis. (J) *ST3GAL3* expression in human PBMCs across publicly available flow-sorted or single-cell RNA-seq datasets.

To assess the relationship between highly correlated gene expression changes (Δ) during the vaccination period and outcomes, we constructed data-driven modules for 230 infants with paired transcriptomes using WGCNA and correlated them to parasitemia and CSP-specific IgG variables. Six modules positively correlated with either protection or days to first parasitemia, and a single *KDM5D*-led module negatively correlated with protection (**Figure 4E**). The protective modules form two distinct networks, one driven by *CD22*, which encodes an inhibitory coreceptor of the B-cell receptor and *BTLA*, which encodes B- and T-lymphocyte-associated protein, and the other driven by *RIOK3, SEC62*, and *TRIP12*, which encodes for an E3 ubiquitin-protein ligase (**Figure 4F**). Modules hubbed by *RIOK3, SEC62*, and *TRIP12* also negatively correlated with both pre-immunization CSP-specific IgG and the number of Pf infections during the vaccination period (**Figure 4E**), suggesting that Pf exposure also negatively impacts protection associated with these modules. By enrichment analysis, *BTLA-* and *CD22*-led modules are predominantly related to B cells and plasma cells and, to a lesser extent, immune activation and inflammation, whereas the *RIOK3*-, *SEC62*-, and *TRIP12*-led modules are related to erythroid cells and heme metabolism (**Figure S5B**). The orphan protective module led by *NKG2E* was enriched in genes related to cytotoxic lymphocytes, and the *KDM5D*-led module was enriched in Y chromosome-linked genes (**Figure S5B**). The data-driven network analysis demonstrates changes in networks related to B cells, cytotoxic lymphocytes, and erythroid cells correlate with protection and provides evidence of an interaction between Pf parasitemia and protective immune responses.

Specifically for the 1.8×10^6^ PfSPZ vaccinated infants, paired comparisons of post-immunization versus baseline transcriptomes by DGE within the protected (ΔP) or not protected (ΔNP) outcomes revealed predominantly negative enrichment of innate and adaptive modules using high-level BTMs, with the exception of NK and T cell modules within ΔNP, which showed positive enrichment (**Figure 4G**). This resulted in significant downregulation of antigen presentation, cell cycle, monocyte, NK cell and T cell modules in ΔP relative to ΔNP (**Figure 4G**). Using Monaco modules, B cell, plasmablast, and naive CD4^+^ T cell signatures are significantly induced after vaccination in ΔP relative to ΔNP. By contrast, cytotoxic lymphocyte signatures are significantly induced after vaccination in ΔNP relative to ΔP (**Figure 4G**), differences that could reflect trafficking of these cell subsets to and from the peripheral blood. Among 6 genes that met 3 criteria of being significantly different between in ΔP versus ΔNP (p<0.005), induced in ΔP (p<0.05), and decreased in ΔNP (p<0.10) post-immunization within 1.8×10^6^ PfSPZ (**Figure 4H; Table S3**), the antisense transcript *RP3-330M21*.*5* and *ST3GAL3* were identified as being associated with reduced risk of parasitemia when applied to infants across all PfSPZ Vaccine dose groups, even when adjusted for age, site, and Pf infections during the vaccination period (**Figure 4I**; **Table S4**). *ST3GAL3* encodes for a sialyltransferase and, based on public flow-sorted and single-cell RNA-seq datasets, is predominantly expressed in CD8^+^ and regulatory T cells in the peripheral blood (**Figure 4J**). Overall, the paired Δ transcriptomic analyses provide further evidence that humoral and cytotoxic immune responses play a role in protection from Pf parasitemia.

### Integrated multi-modal analyses reveal features predictive of PfSPZ-induced protection from parasitemia

We integrated gene expression data with cellular expression data, including immunophenotyping of rested and PfSPZ-stimulated PBMCs and frequency of CSP-specific B cells, and plasma cytokines to assess their linear relationships with each other and with outcomes of post-immunization CSP-specific IgG and time to parasitemia up to 6 months. To reduce the number of features, gene expression was collapsed to module expression scores using either high-level BTMs or Monaco modules. Among baseline features, only the NK and mucosal-associated invariant T (MAIT) cell modules positively correlated with time to parasitemia, and only plasma TNF positively correlated with post-immunization CSP-specific IgG (FDR<5%; **Figure 5A; Table S5**). Notably, at baseline, peripheral CD14^+^CD16^+^ ‘non-classical’ monocytes positively correlated with the plasma cytokines CXCL10/IP-10, IFNα, and IL-10 as well as with peripheral plasmablasts (**Figure 5A**). Using post-immunization data, CSP-specific memory B cells, IgG+ of CSP-specific B cells, and CSP-specific IgG correlated with time to parasitemia, consistent with above (**Figures S6A and 3A; Table S5**). CSP-specific memory B cells also correlated with the Monaco module related to basophils, which have been shown to enhance humoral memory responses (Denzel et al., 2008). Non-classical CD14^low^CD16^+^ monocytes positive correlated with the Monaco cytotoxic modules memory CD8^+^ T, NK, Vδ2 γδ T, and non-Vδ2 γδ T cells (**Figure S6A; Table S5**). Time to parasitemia did not significantly correlate with any features when expressed as the Δ (log_2_ fold-change of post-immunization over pre-immunization baseline values; **Figure S6B; Table S5**). Changes in frequency of atypical memory B cells, which expand after PfSPZ vaccination in malaria-naïve donors (Sutton et al., 2021), positively correlated with changes in high-level BTMs related to cell cycle, DC activation, IFN/antiviral sensing, and signal transduction (**Figure S6B; Table S5**).

**Figure 5.**
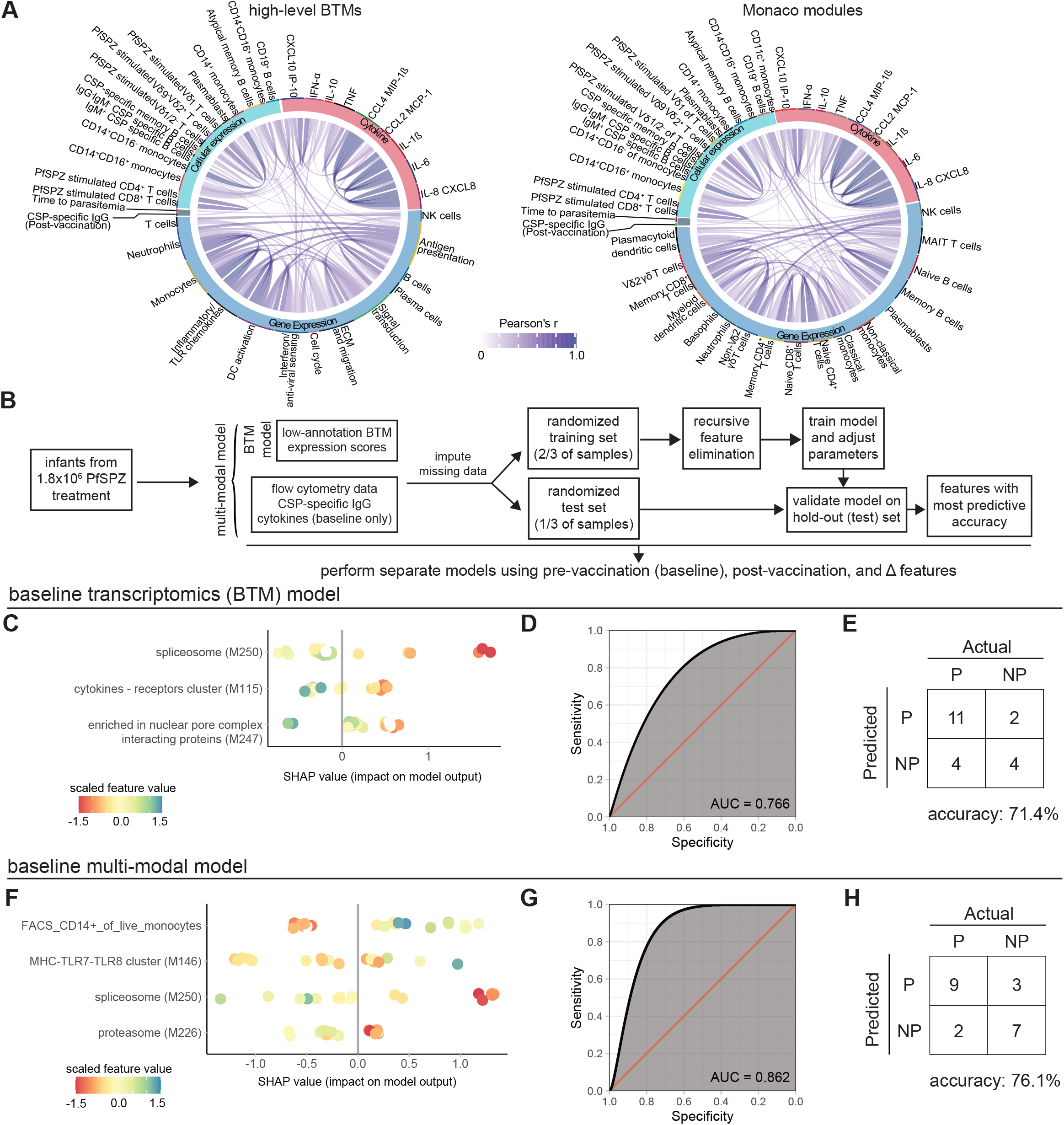
Integrated multi-modal analyses reveal features predictive of PfSPZ-induced protection from parasitemia. (A) Pairwise correlations between baseline features (gene expression collapsed into either high-annotation level blood transcription or Monaco modules, flow cytometric features, and plasma cytokines) with the post-vaccination outcomes of plasma CSP-specific IgG and time-to-parasitemia at 6 months for all infants with available data. Blue lines indicate significantly positive correlations (FDR<0.20). (B) Overview of machine learning workflow. XGBoost was used to predict the outcome of parasitemia at 3 months using a blood transcriptomic module (BTM) model or a multi-modal model that combined BTM features with flow-cytometric, CSP-specific IgG, and cytokine features. (C) SHapley Additive exPlanations (SHAP) plots for BTM model showing marginal contribution of each feature to the predicted outcome based on feature values. Each point is a single subject in the test set. (D) Receiver operating characteristics (ROC) curve for BTM model. AUC = area under the curve. (E) Confusion matrix showing the model predictions and actual outcomes in the test set for BTM model. (F) SHAP plot, (G) ROC curve, and (H) confusion matrix for the multi-modal model.

We hypothesized that a non-linear machine learning approach may better identify features predictive of the binary outcome of P or NP for the 1.8×10^6^ PfSPZ group. Using a gradient boosted decision tree algorithm, we built models that included only low-annotation BTM expression scores and multi-modal models that added additional features, with separate models performed for baseline, post-immunization, and Δ datasets (**Figure 5B; Table S6**). The baseline transcriptomics model identified the “spliceosome” (M250), “cytokines-receptors cluster” (M115), and “enriched in nuclear pore complex interacting proteins” (M247) as the top features with an area under the curve (AUC) of 0.633 and a predictive accuracy of 71.4% when applied to the test set (**Figure 5C-E**). The better performing baseline multi-modal model identified CD14^+^ monocytes, the antigen processing and presentation “MHC-TLR7-TLR8 cluster” module (M146), “spliceosome” (M250), and “proteasome” (M226) as the top features with an AUC of 0.862 and a predictive accuracy of 76.1% when applied to the test set (**Figure 5F-H**). Notably, lower feature values for CD14+ monocytes and M146 were predicted to have higher impact on protective outcomes (**Figure 5F**), consistent with our data above showing that innate immune activation signatures were enriched at baseline in NP infants in the 1.8×10^6^ PfSPZ group (**Figure 2D-E**). The post-immunization models identified “leukocyte migration” (M88.0), “putative SREBF1 targets” (M178), and “cell adhesion, membrane” (M133.0) as features predictive of protection (**Figure S6C**), while “CORO1A-DEF6 network (II)” (M32.4), “plasma cells, immunoglobulins” (M156.1), “cell cycle, ATP binding” (M144), and “complement activation (I)” (M122.0) were top features in the Δ models **(Figure S6D**). However, none of these models performed as accurately as the multimodal baseline model (**Table S6**).

### Innate immune activation enhances monocyte phagocytic capacity of sporozoites independent of CSP-specific antibodies

Protective correlation of a baseline gene network related to Fcγ receptor-mediated phagocytosis (**Figure 2C**), higher baseline CSP-specific IgG in NP infants (**Figure 3D**), and the identification of CD14^+^ monocytes as the top baseline predictor of protection (**Figure 5F**) within the 1.8×10^6^ PfSPZ group prompted us to examine these variables in the context of treatment and protection. We observed a reciprocal interaction between CD14^+^ monocytes and CSP-specific IgG at baseline (**Figure 6A**) in which, for infants with CSP-specific IgG>0, CD14+ monocyte percentage is higher in P versus NP for placebo but lower in P versus NP for 1.8×10^6^ PfSPZ (**Figure S7A**). The treatment-dependent effect of CSP-specific IgG and increased peripheral CD14+ monocytes at baseline on protective outcome suggests that Ab-dependent opsonic phagocytosis may provide another mechanism that confers short-term protection to natural infection while preventing protection by PfSPZ Vaccine. We revisited the observation that innate immune activation had a similar treatment-dependent effect on outcome (**Figure 2E**) and asked if differential innate activation could be seen in infants independent of pre-existing CSP-specific IgG. Among 1.8×10^6^ PfSPZ infants lacking baseline CSP-specific IgG, innate activation signatures are generally higher in NP versus P, with significance observed for modules related to host antiviral responses (M111.0, M111.1, M150), IFN (M75, M127), antigen presentation (M5.0, M95.0), “activated DCs” (M66), “inflammasome receptors and signaling” (M53), “MHC-TLR7-TLR8 cluster” (M146), and “lysosome” (M209) (**Figure 6B; Table S7**). Notably, “MHC-TLR7-TLR8 cluster” was a top predictor in the baseline multi-modal model (**Figure 5F**), and lysosome-related genes may represent a pan-innate immune memory signature (Jentho et al., 2019). Conversely, for 1.8×10^6^ PfSPZ infants with CSP-specific IgG at baseline, expression of the antigen presentation and lysosome modules (M5.0, M95.0, M209) was increased in P versus NP (**Table S7**), consistent with baseline enrichment of the antigen presentation genes in high-CSP IgG responders, which generally correlates to protection (**Figure 3D-E**). No significant differences in module expression were observed between outcomes for placebo with or without CSP-specific IgG (**Table S7**). Given that pre-activation with the TLR4 agonist LPS can augment the phagocytic efficiency of *Plasmodium*-infected erythrocytes by liver macrophages (Ono et al., 2021), we hypothesized that innate activation can also enhance the capacity of monocytes to phagocytose sporozoites in an Ab-independent manner. Pre-treatment with either LPS or the TLR5 agonist flagellin, both which signal via myD88 (Gewirtz et al., 2001; Kawai et al., 2001), significantly increased the capacity of human monocytes to phagocytose live *P. yoelii* (*Py*, a rodent malaria parasite) sporozoites relative to vehicle (**Figure 6C-E; Figure S7B**). In contrast, the TLR9 agonist CpG and β-glucan, a dectin-1 agonist that acts via a non-TLR signaling pathway (Gross et al., 2006), significantly decreased monocytic phagocytosis. The TLR3 agonist poly I:C and TLR7 agonist imiquimod had no significant effect on phagocytosis. These data suggest that opsonin-independent phagocytosis can be modulated by specific innate microbial signals.

**Figure 6.**
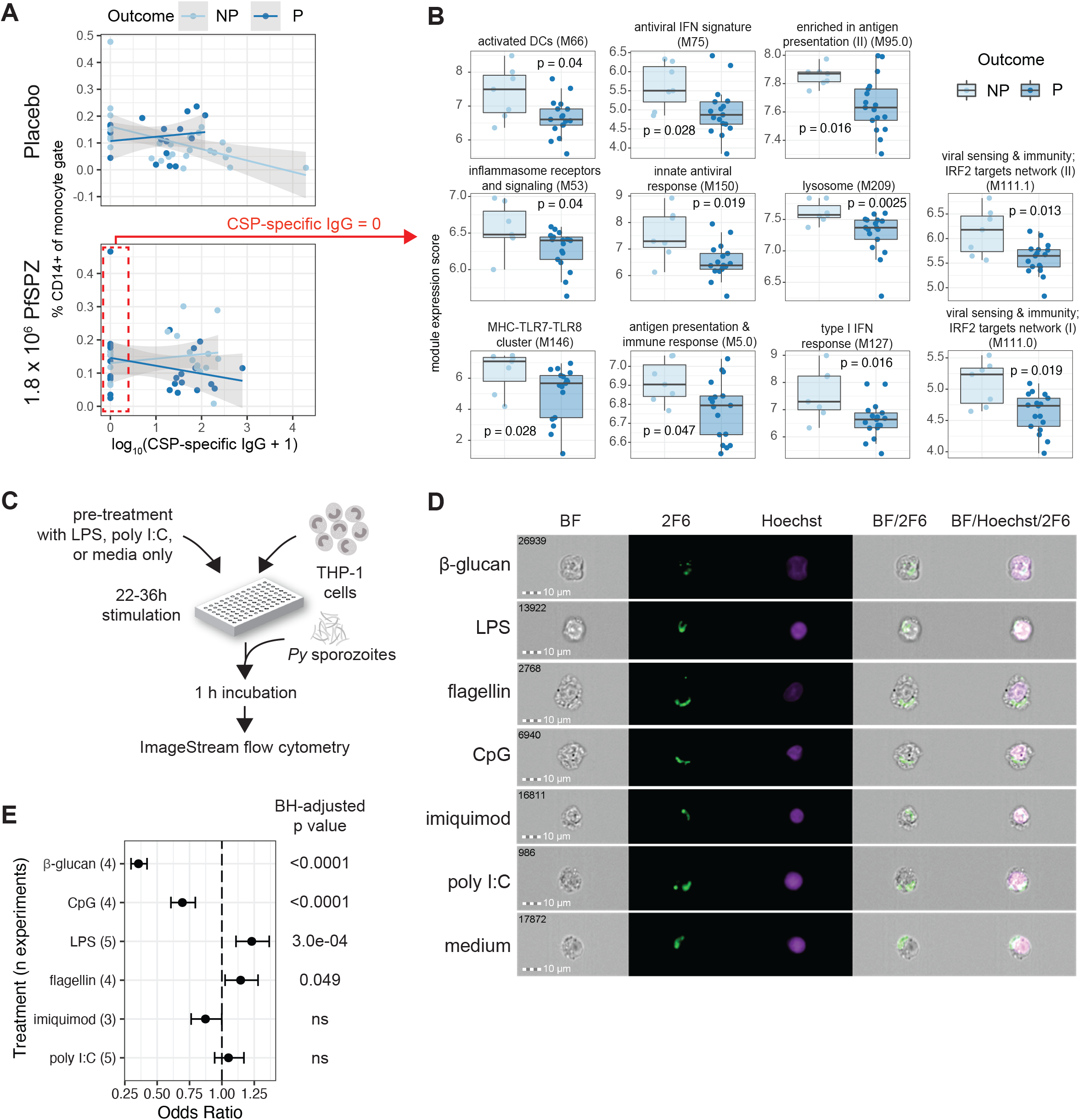
Innate immune activation enhances monocyte phagocytic capacity of sporozoites independent of CSP-specific antibodies. (A) Interaction between % CD14+ peripheral monocytes and CSP-specific IgG at pre-immunization baseline and protective outcome differs between placebo and 1.8×10^6^ PfSPZ groups. (B) Pre-immunization baseline expression of innate-related low-annotation BTMs in non-protected (NP) and protected (P) infants who received 1.8×10^6^ PfSPZ and lacked baseline CSP-specific IgG. (C) Schematic for *in vitro* sporozoite phagocytosis assays. (D) Representative images of *P. yoelii* (Py) sporozoites stained using anti-PyCSP 2F6 monoclonal Ab after pre-treatment with indicated conditions. BF = brightfield. (E) Odds ratios with 95% confidence intervals for number of THP-1 cells containing phagocytosed *Py* sporozoites over all THP-1 cells for indicated treatment versus medium only control across indicated number (n) of independent experiments. Significance determined by Fisher’s exact test.

### Stimulation of innate immunity can confer protection against liver parasite burden but dampens RAS-induced CD8^+^ T cell responses

Prior studies have shown that innate immune activation can inhibit *Plasmodium* liver-stage development (Chen et al., 2009; Gramzinski et al., 2001; Liehl et al., 2014) and protective adaptive immune responses against the pre-erythrocytic stage (Minkah et al., 2019; Murphy et al., 2021). Given the increased innate activation observed prior to immunization in infants who became parasitemic after receipt of 1.8×10^6^ PfSPZ Vaccine (**Figure 2D-E**), we tested the hypothesis that innate stimulation inhibits CD8^+^ T cell priming by reducing liver-stage burden using the *Py* 17XNL RAS immunization model in malaria-naive C57BL/6 mice. Pre-treatment with either LPS or poly I:C, but neither flagellin nor β-glucan, reduced *Py* liver-stage burden after non-irradiated, fully infectious *Py* sporozoite injection when compared to saline control (**Figure 7A-B**), consistent with a prior study (Chen et al., 2009). Pre-treatment with either flagellin, LPS, or poly I:C, but not β-glucan, dampened the RAS-induced increase in circulating antigen-experienced CD11a^hi^CD8^lo^ T cells (Rai et al., 2009) 7 to 28 days post immunization (**Figure 7C-F; Figure S7C**). Taken together, this data suggests that pre-existing activation of specific innate signaling pathways can reduce priming of antigen-specific CD8^+^ T cells by RAS by limiting liver-stage infection.

**Figure 7.**
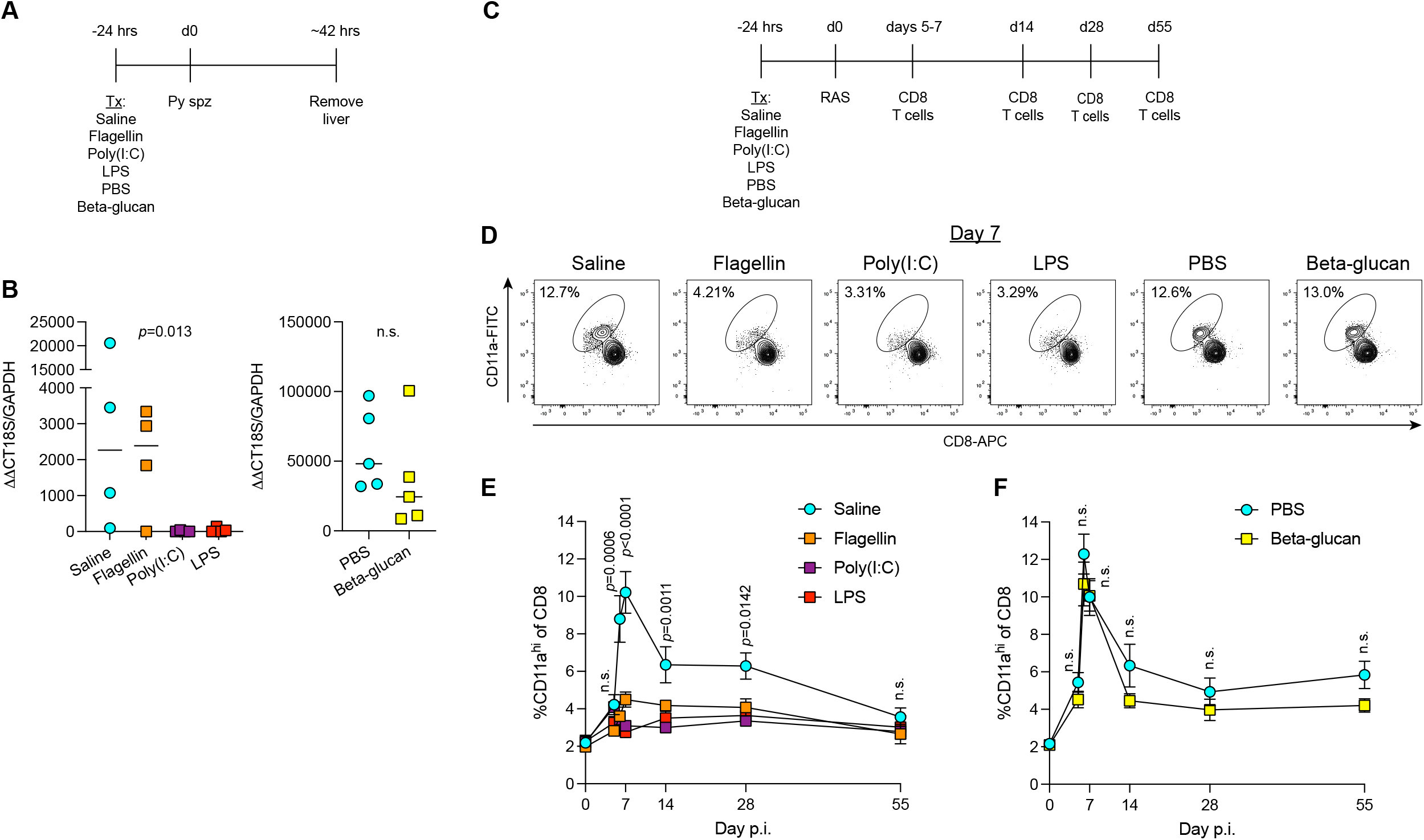
Stimulation of innate immunity can confer protection against liver parasite burden but dampens RAS-induced CD8^+^ T cell responses. (A) C57BL/6 mice were treated with the indicated innate stimuli or vehicle control (saline or PBS) 24 h before injection of 1000 *P. yoelii* (*Py*) 17XNL sporozoites. Liver tissue was removed ∼42 h after sporozoite injection. (B) Quantification of liver parasite burden. Each symbol represents a single mouse. Data (median) are representative of two independent experiments were analyzed by Kruskal-Wallis test, n.s. = not significant. (C) C57BL/6 mice were treated with the indicated innate stimuli or vehicle control (saline or PBS) 24 h before injection of about 1×10^4^ radiation attenuated *Py* 17XNL sporozoites (RAS). RAS-induced CD8^+^ T-cell responses were enumerated in peripheral blood on the indicated days. (D) Representative flow cytometry plots identifying RAS-induced CD8^+^ T cell (CD8^lo^CD11a^hi^). Value within plot is the percent of all circulating CD8^+^ T cells that are CD8^lo^CD11a^hi^. (E) Percent of all circulating CD8^+^ T cells that are CD8^lo^CD11a^hi^ on the indicated day post-RAS injection. Data (mean±S.E.) are cumulative results (n=8 mice/treatment) from two independent experiments and were analyzed by Kruskal-Wallis test, n.s. = not significant. (F) Percent of all circulating CD8^+^ T cells that are CD8^lo^CD11a^hi^ on the indicated day post-RAS injection. Data (mean±S.E.) are cumulative results (n=9-10 mice/treatment) from two independent experiments and were analyzed by Mann-Whitney test, n.s. = not significant.

## DISCUSSION

The efficacy and durability of protective vaccine responses can be profoundly influenced by host intrinsic (e.g. age, gender, and genetics) and extrinsic (i.e. pre-existing immunity, microbiota, prior infections, co-infections) factors (Zimmermann and Curtis, 2019). Systems analyses of vaccination regimens can elucidate the early immunological processes that drive pathogen-specific adaptive responses and protective efficacy to help identify these factors and inform rational vaccine design (Fourati et al., 2016; Hagan et al., 2019; Kotliarov et al., 2020; Li et al., 2017; Nakaya et al., 2011; Tsang et al., 2014). Here, we provide a comprehensive systems analysis of a clinical trial of the radiation-attenuated PfSPZ Vaccine conducted in infants living in the high malaria transmission setting of western Kenya that identified baseline innate immune activation as a key differentiator of vaccine response.

Several observations from the current analysis are in line with what is known about sterile immunity to Pf. Baseline enrichment of genes related to NK, γΔ T, and CD8^+^ T cells in protected versus non-protected infants for both the placebo and 1.8×10^6^ PfSPZ groups (**Figure 2D**) may reflect shared cytotoxic gene signatures and also is consistent with prior studies demonstrating the protective roles of each of these cell types against liver-stage infection (Epstein et al., 2011; Jagannathan et al., 2017; Roland et al., 2006; Zaidi et al., 2017). The correlation of post-vaccination CSP-specific IgG and memory B cells with subsequent protection (**Figure 3A-B**) is also consistent with the protective role of neutralizing CSP-specific Abs against Pf sporozoites (Dobano et al., 2019; Gaudinski et al., 2021; Kisalu et al., 2018; Tan et al., 2021; Tan et al., 2018; Wang et al., 2020; White et al., 2013). Additionally, despite the lack of Pf-specific cellular responses to PfSPZ Vaccine in this infant cohort (Oneko et al., 2021), we did observe significant changes in adaptive modules (including induction of memory B cells, plasmablasts, and plasma cells) after high-dose PfSPZ vaccination that correlated with protection, perhaps reflecting expansion or activation of Pf-specific cells.

Our analysis also revealed additional insight into innate immunity against Pf infection in malaria-exposed infants. We previously suggested that the lack of T-cell responses in infants in this trial may be associated with the relative lack of VΔ2 T cells in this age cohort, which can activate and expand in response to phosphoantigens contained in sporozoites, at the time of first immunization (Oneko et al., 2021). The current study provides evidence for an alternative mechanism involving innate activation of monocytes and possibly other phagocytic cells. Reciprocal enrichment of myeloid and innate inflammatory signatures in the protected infants within the placebo group and in unprotected infants within the 1.8×10^6^ PfSPZ (**Figures 2 D-E, Figure S3**) suggests that innate immune activation may confer short-term protection against natural, fully infectious sporozoites but also may prevent the liver-stage infection necessary for PfSPZ Vaccine to generate effective CD8^+^ T cell responses and achieve durable protection (Epstein et al., 2011; Ishizuka et al., 2016).

We provide evidence that, in the absence of sporozoite-specific IgG, pre-stimulation of TLR pathways can enhance phagocytosis of sporozoites *in vitro* and restrict priming of antigen-specific CD8^+^ T cells after RAS inoculation by reducing liver-stage burden *in vivo*. Notably, LPS, a TLR4 agonist that acts via both myD88- and TRIF-dependent innate signaling pathways (Kawai et al., 2001; Yamamoto et al., 2003), enhanced sporozoite phagocytosis, decreased *Py* liver-stage burden, and restricted CD8^+^ T cell priming by RAS. In contrast, poly I:C, a TLR3 agonist that signals solely through TRIF (Yamamoto et al., 2003), decreased liver-stage burden and CD8^+^ priming but did not enhance phagocytosis. Thus, innate activation may restrict CD8+ priming by acting directly on hepatocytes via TRIF-dependent pathways to reduce RAS liver-stage progression. The innate activation observed in the peripheral blood of infants may reflect systemic activation that directly affects highly phagocytic tissue-resident macrophages (Gordon, 2016) or hepatocytes, in which IFN has been shown to reduce liver-stage replication and restrict protective adaptive immune responses against sporozoites (Liehl et al., 2014; Miller et al., 2014; Minkah et al., 2019). Our findings differ from a recent transcriptional analysis of Pf-stimulated PBMCs in young African children immunized with RTS,S/AS01E in which innate, inflammatory gene signatures both pre- and post-immunization predicted protection from malaria (Moncunill et al., 2020). However, in the current study, baseline innate activation was also associated with enhanced CSP-specific IgG responses induced by PfSPZ vaccination (**Figure 3C**), consistent with prior evidence that triggering TLRs on antigen-presenting cells during immunization can greatly enhance Ab responses (Kasturi et al., 2011; Oh et al., 2014). Taken together, these findings uncouple protective immunity achieved by antibodies from cytotoxic responses and suggest that the efficacy of PfSPZ Vaccine in malaria-endemic settings might be constrained by opposing antigen presentation pathways.

Our data suggests that multiple innate triggers, with the notable exception of β-glucan, may be capable of inhibiting efficacy of the PfSPZ Vaccine. In a malaria-intense setting, a likely culprit is Pf parasitemia. Indeed, *Plasmodium*-infected erythrocytes can activate macrophages via the NLRP3 and AIM2 inflammasomes (Kalantari et al., 2014). The appearance of heme- and erythroid-related signatures before and after the vaccination period provides indirect evidence for recent or sub-microscopic Pf parasitemia. At baseline, the erythropoiesis-related gene *WDR26* (Zhen et al., 2020) was a strong driver of variation, and heme and erythroid signatures positively correlated with pre-existing CSP-specific IgG and Pf parasitemia at first immunization and negatively associated with protection (**Figures 2A-C and S3C**). Such erythroid signatures were detected after effective globin depletion (**see Methods**) and likely represent reticulocyte transcripts given the gradual loss of RNA as erythrocytes mature. A negative correlation with malaria protection can be interpreted in epidemiological terms in that recent malaria, resulting in compensatory erythropoiesis, also predicts future incidence of parasitemia, which is supported by the enrichment of erythroid genes in unprotected infants across all treatment groups (**Figure S3C**). However, it is also possible that low-level Pf parasitemia reduces protective responses to PfSPZ Vaccine. The negative impact of parasitemia on whole-sporozoite immunization is supported by a study showing that Pf blood-stage parasitemia decreases the efficacy of PfSPZ-CVac, an approach which involves immunization of fully infectious PfSPZ under the cover of malaria chemoprophylaxis (Murphy et al., 2021).

## LIMITATIONS OF THE STUDY

We did not assess for sub-microscopic Pf parasitemia, co-infections with other pathogens during the vaccination period, or differences in intestinal microbiota, which may have aided the identification of specific microbial triggers of innate immunity. Given the practical limits for volume and frequency of blood collections in this field trial of infants, our transcriptomic analysis relied on bulk RNA-seq of *ex vivo* whole blood sampled at only two time points. Thus, we could not differentiate activation from proportional differences in cell types, and having a single post-immunization time point reduced our sensitivity for detecting Pf-specific cellular responses. Future work will be needed to assess the differential kinetics between the blood transcriptomes of vaccine-responders and non-responders at the single-cell level. We did not directly test whether Pf-infected erythrocytes could enhance monocyte phagocytic capacity of Pf sporozoites. Lastly, as the trial was conducted in a high malaria transmission setting, our findings may not be generalizable to areas with less intense malaria transmission.

## CONCLUSIONS

In summary, we present evidence supporting a model whereby baseline innate immune activation is associated with short-term protection from natural *P. falciparum* infection but also reduced liver-stage infectivity by the radiation-attenuated PfSPZ malaria vaccine which restricts priming of antigen-specific CD8^+^ T cell responses and protective efficacy. Our study identified states of innate immune activation that could potentially be modulated by drug treatment to improve the efficacy of whole-sporozoite immunization regimens. Alternatively, screening for innate immune activation prior to vaccination could identify those who are the mostly likely to respond to whole-sporozoite malaria vaccine regimens.

## METHODS

### LEAD CONTACT AND MATERIALS AVAILABILITY

Requests for further information and resources or reagents should be directed to and will be fulfilled by the lead contact Tuan M. Tran.

## EXPERIMENTAL MODELS AND SUBJECT DETAILS

### Human studies

Details of the KSPZV1 clinical trial have been described (Oneko et al., 2021; Steinhardt et al., 2020). Briefly, the trial was conducted from January 2017 to August 2018 in Siaya County, western Kenya, where malaria transmission is highly intense and occurs year-round with peaks during the long (April–July) and short (October–November) rainy seasons. Infants aged 5 to 12 months were randomized to receive PfSPZ Vaccine dosages of 4.5×10^5^, 9.0 ×10^5^, 1.8×10^6^ PfSPZ or normal saline placebo in a 1:1:1:1 ratio. Three vaccinations or placebo injections were administered intravenously at 8-week intervals. Artemisinin-based combination therapy (ACT), primarily artemether-lumefantrine, was administered to all study participants 11-19 days prior to the last vaccination to clear parasitemia at the beginning of active and passive surveillance for clinical malaria and *Pf* infection. Cases of *Pf* infection were determined by active surveillance at scheduled monthly, in case of symptoms during scheduled interim visits, and by passive surveillance during symptom-triggered clinic visits, during which a rapid diagnostic test (RDT) or contemporaneous blood smear was performed. Blood smears were prepared for subsequent reading, and dried blood spots on filter paper (DBS) were collected for subsequent molecular testing using polymerase chain reaction (PCR) during active surveillance visits and during interim or sick visits if children were febrile (axillary temperature ≥37.5°C or reported fever in the last 24 h); febrile children were treated for malaria according to rapid diagnostic test or expedited microscopy results. For the current study, the primary outcome was presence of *Pf* parasitemia (not protected; NP) or absence of *Pf* parasitemia (protected; P), as determined by microscopy, through 3 months of surveillance post-immunization. We also used a secondary outcome of time (days) to first *Pf* parasitemia up to 168 days (∼6 months) post-immunization.

#### Ethics statement

Written informed consent was obtained from a parent/guardian of each infant. The clinical study protocol was approved by institutional review boards (IRB) of the Kenya Medical Research Institute (KEMRI), the US-based Centers for Disease Control and Prevention (CDC), the Kenya Pharmacy and Poisons Board (PPB) and was registered at ClinicalTrials.gov (NCT02687373). The laboratory study protocol for secondary use of de-identified human samples, including RNA sequencing, was approved by the Indiana University IRB.

### Mouse studies

Six-week-old female C57BL/6 mice were purchased from Charles River Labs. Daily care was provided by Laboratory Animal Resource Center (LARC) at Indiana University School of Medicine (IUSM). Seven-week-old female C57BL/6 mice were treated intravenously with either saline (0.9%; Teknova, Cat. S5825), flagellin (10 μg; Adipogen, Cat. AG-40B-0095-C100), poly I:C (200 μg; Tocris, Cat. 4287), or lipopolysaccharide (LPS; 10 μg; Sigma, Cat. L3024-5MG) or via intraperitoneal injection with endotoxin-free phosphate-buffered saline (PBS; Corning, Cat. 21-040-CV) or β-glucan (1 mg; Sigma, Cat. G5011-25MG) 24 h prior to immunization. Radiation attenuated sporozoites (RAS) were diluted to 5×10^4^/mL in sterile saline; each mouse received 1×10^4^ RAS intravenously and untreated mice received 10^3^ viable sporozoites intravenously. All injections were in 200 μL volume.

#### Ethics statement

Approval for the animal studies was obtained from IUSM Institutional Animal Care and Use Committee (IACUC) under protocol 19024 in compliance with all applicable federal regulations and accredited by AAALAC, International.

### Sporozoite isolation

*Anopheles stephensi* mosquitoes infected with *P. yoelli* were purchased from the Insectary at Seattle Children’s Research Institute. Salivary glands were dissected into Roswell Park Memorial Institute 1640 Medium (RPMI, Gibco, Cat. 11875-093) and sporozoites were isolated using the Ozaki protocol (Ozaki et al., 1984). Irradiation of the sporozoites was performed using the following parameters: 200 Gy; ∼519 cGy/min for 38.5 min.

## METHOD DETAILS

### Human studies

#### Sample collection

Blood samples were collected by venipuncture in PAXgene Blood RNA (BD Diagnostics), Vacutainer serum separator (Becton-Dickinson), and Vacutainer K2EDTA (Becton-Dickinson) tubes and labeled, stored, and shipped in line with good clinical and laboratory practice principles. Drops of capillary blood were used to make thick blood smears and dried blood spots (DBS) on filter paper.

#### Malaria detection by microscopy

Blood slides were prepared at all scheduled visits and at unscheduled visits where a malaria test was indicated due to fever or history of fever in the last 24 h. Blood smears were not read in real-time unless children had fever or history of fever, in which case the smear was read immediately, and children treated if positive. Malaria infection and the parasite densities were determined by two certified readers. In the case of discordant results (positive/negative discordancy; greater than 2-fold discrepancy if parasitemia >400 parasites/µL; greater than 10-fold discrepancy if parasitemia < 400 parasites/ µL), a third read was carried out.

### RNA processing and sequencing

#### KSPZV1 study

RNA extraction and sequencing for all subjects were performed in 96-well plate format as two separate batches of four and two plates, in which subjects were randomized in a treatment-stratified manner to each plate. Paired pre- and post-vaccination samples from the same subject were on the same plate. Total RNA was extracted from PAXgene stabilized whole blood using the PAXgene 96 Blood RNA kit (Qiagen) and treated with RNase-Free DNase Set (Qiagen). RNA quality was assessed by automated parallel capillary electrophoresis on a Fragment Analyzer (Advanced Analytical/Agilent). The average RQN was 8.4. For each sample, 100 ng of total RNA was used for library preparation using Biomek FXP automation for 96 well plates. Ribosomal and globin mRNA were removed using QIAseq FastSelect rRNA and QIAseq FastSelect GlobinRNA removal kit, respectively (Qiagen, human). RNA was fragmented, converted to cDNA, ligated to index adaptors, and amplified using the KAPA RNA HyperPrep Kit (Roche). Quantification and quality were assessed again using Agilent TapeStation and libraries were pooled with QIAgility (Qiagen). RNA sequencing of 150 bp paired-end reads were performed on the NovaSeq 6000 Sequencing System v1.0 (Illumina). Illumina sequences were trimmed of contaminating adapters and bases. After assessing sequencing quality using FastQC (v0.11.5, Babraham Bioinformatics, Cambridge, UK), paired-end reads meeting a Phred quality score (Q score) > Q30 were mapped to reference human genome GRCh38 (version 16, Ensembl 99) using STAR RENA-seq aligner (v2.5). The following parameter was used for mapping: “Se--outSAMmapqUnique 60”. Assessment of reads distribution was performed using bamutils (ngsutils v0.5.9). Uniquely mapped reads were assigned to hg38 refGene genes using featureCounts (subread v1.5.1) with parameters “-s 2 –p –Q 10”. After sequencing genomic DNA (gDNA) contamination was suspected due to high intergenic read percentages for some samples. The SeqMonk RNA-Seq quantitation pipeline was used to correct for gDNA contamination using the FASTQ results from Illumina sequencing (Babraham Bioinformatics, Cambridge, UK). Expression of 88 genes encoding select lineage markers or relevant to immune responses was validated using nCounter PlexSet (nanoString), with 57 genes (65%) exhibiting strong correlation (Spearman ρ≥0.6) between RNA-seq and nCounter expression values.

#### VRC studies

RNA was extracted from PAXgene blood RNA tubes using QIAGEN Rneasy kit with on-column Dnase digestion and steps to remove rRNA and globin RNA contamination from erythrocytes, according to manufacturer’s instructions. First-strand Illumina-barcoded libraries were generated using the NEBNext Ultra Directional RNA Library Prep Kit for Illumina, using the mRNA capture kit and 12-16 cycles of PCR enrichment, according to manufacturer’s instructions. Stranded libraries were sequenced on an Illumina HiSeq 2500 instrument using paired-end 50-bp reads. Data were trimmed for quality using Trimmomatic v0.36 with the following parameters: LEADING:15 TRAILING:15 SLIDINGWINDOW:4:15 MINLEN:37. Trimmed reads were aligned to the hg19 human genome assembly using Bowtie2 v2.2.9, and reads were quantified using HTSeq v0.9.1.

### Differential gene expression analysis

#### KSPV1 study

Differential gene expression (DGE) analysis was performed using edgeR v.3.30.3 (Robinson et al., 2010) to compare subjects who would later remain uninfected (protected; P) or become parasitemic with *P. falciparum* (not protected; NP) by 3 months after the last vaccination. Lowly expressed genes were filtered using the filterByExpr function using group sizes defined by treatment-outcome-timepoint parameterization, and the remaining genes were normalized using the weighted trimmed mean of M-values (TMM) method (Robinson and Oshlack, 2010). Samples with mapped library sizes <7.5 million counts were excluded. Normalized counts were used for expression analysis. For the baseline analysis, filtering and normalization were performed separately for samples within each treatment group. After gene-specific dispersion estimation, DGE between 3-month outcomes was determined using the glmQLFtest function and the following model matrix formula and contrast:

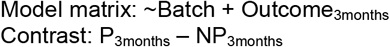

Analysis of post-vaccination samples was performed as a paired analysis with each subject serving as their own baseline control. No adjustments for batch were performed given that paired samples for each subject were performed on the same plates for RNA extraction and sequencing. Thus, for DGE between the post-vaccination and baseline timepoints *within* an outcome group, the following model matrix and contrast were used:

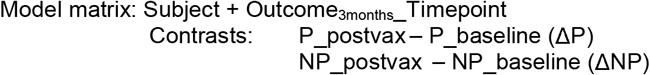

Here Outcome_3months__Timepoint represents the combined parameterization of outcome and timepoint with baseline as the reference level. To compare the post-vaccination effect between outcome groups while accounting for baseline, the following contrast was used:

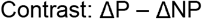

Where Δ denotes post-vaccination – baseline within each outcome group.

#### VRC datasets

For the VRC 312 and VRC 314 datasets, only baseline transcriptomes were analyzed. Both of these clinical trials contained multiple PfSPZ Vaccine dosing regimens (Ishizuka et al., 2016; Seder et al., 2013). In the current study, analysis was limited to regimens with a constant PfSPZ Vaccine dose delivered multiple times intravenously (**Figure S4A**). Analysis was similar as the baseline analysis for the KSPZV1 trial but with additional adjustment for dosing regimen:

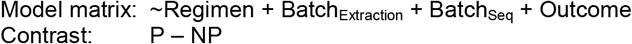

#### Enrichment and regulator analysis

Gene set enrichment analysis (GSEA) (Subramanian et al., 2005) was performed with fast GSEA (Korotkevich et al., 2021) using the fgseaMultilevel function on genes ranked by - log_10_(p value)*sign(log_2_ fold-change), with significance and fold-change values obtained from the DGE analyses. Minimum gene set size was set to 20 and gene sets used were low-annotation blood transcription modules (BTMs) (Li et al., 2014), high-annotation BTMs (Kazmin et al., 2017), BloodGen3 modules (Rinchai et al., 2021), or modules derived from the Monaco et al. RNA-seq dataset (Monaco et al., 2019). For the Monaco modules, a gene was included within a cell type-specific module if its expression z-score (scaled across all cell types) was greater than 1.75 for that cell type. Additional GSEA was performed using the Hallmark collection from MSigDB (Liberzon et al., 2015) and KEGG pathway database (Kanehisa et al., 2016). Upstream regulator analysis was performed using genes with a |log_2_ fold-change|>1.2 and p<0.25 as determined by DGE above in Ingenuity Pathway Analyis (Qiagen, March 2021 release) (Kramer et al., 2014).

### Weighted gene correlation network analysis

Weighted gene correlation network analysis (WGCNA) was performed using normalized counts per million (log_2_CPM) for at baseline (n=244) and for Δ (log_2_CPM_post-vax_ - log_2_CPM_baseline_, n=230) using the WGCNA package in R (Langfelder and Horvath, 2008, 2012). Using the blockwiseModules function, signed networks were constructed using biweight mid-correlation and soft-thresholding power = 12.5 for baseline analysis and Pearson correlation and soft-thresholding power = 11.5 for Δ analysis. Parameters for module detection included deepSplit=2, minModuleSize=20, mergeCutHeight=0.05, and maxBlockSize set to the total number of genes (single block). Hub genes were identified for networks that significantly correlated with protection at 3 months or time-to-parasitemia through 6 months by Pearson correlation (p<0.05), and these networks were plotted with the igraph package (Csardi and Nepusz, 2006) using parameters optimized to emphasize strong correlations (edges) for better clarity of visualization. To determine biological significance of significant modules, GSEA was applied as above with genes ranked by strength of module membership.

### Survival analysis

Genes meeting the following criteria from the Δ post-vaccination DGE analyses (see above) within the 1.8×10^6^ PfSPZ treatment group were evaluated as potential vaccine-induced predictors of protection against *Pf* parasitemia: (1) significantly different between in ΔP versus ΔNP (p<0.005); (2) significantly induced in ΔP post-vaccination (p<0.05); and downregulated in ΔNP at 2 weeks post-vaccination (p<0.10). For each gene, infants from all PfSPZ Vaccine treatment groups (n=155) were dichotomized as having expression upregulated or downregulated post-vaccination with 1.8×10^6^ PfSPZ if log_2_(CPM_post-vax_/CPM_baseline_) >0 or <0, respectively. Survival analysis was performed using the survival package (Therneau and Grambsch, 2000) in R. Kaplan-Meier curves were used to estimate the probability of remaining free of *Pf* parasitemia during the first 168 days (∼6 months) of surveillance. Log-rank analysis was used to test the significance of differences in time to incident *Pf* parasitemia between infants with or without upregulation of each gene. A Cox proportional hazards model was used to estimate the risk of *Pf* parasitemia between groups and included the following covariates: age (months), site, *Pf* parasitemia during the vaccination period, and PfSPZ Vaccine dose. This combination of variables met the assumptions of proportional hazards. Statistical significance was defined as a 2-tailed□P□value of <0.05.

#### Immunogenicity analysis

Immunogenicity studies were done from blood samples collected at screening and 2 weeks after the third vaccination. Plasma or serum was used for measuring CSP-specific IgG by ELISA as previously described (Mordmuller et al., 2017; Oneko et al., 2021).

#### Flow cytometry analysis

Peripheral blood mononuclear cells (PBMCs) were used to assess the phenotype and function of cellular immune responses by multi-parameter flow cytometry as previously described (Oneko et al., 2021). Immunophenotyping data for T cells was derived from intracellular cytokine stimulation assays previously performed for the parent clinical trial (Oneko et al., 2021) to evaluate T cell responses elicited by PfSPZ Vaccine using previously described methods (Ishizuka et al., 2016). Briefly, cryopreserved PBMCs were thawed using “thawsome” tube adaptors (Beddall et al., 2016) and rested for 8 h, followed by stimulation for 17 h with media control or 1.5 × 10^5^ viable, irradiated, aseptic, cryopreserved PfSPZ from a single production lot (PfSPZ-stimulated). Thus, for T cell subsets, media control samples served as an approximation of ex vivo stained PBMCs. Following stimulation, cells were stained and analyzed as described previously (Jongo et al., 2020). Briefly, cells were washed and stained with viability dye for 20 min at room temperature, followed by surface stain for 20 min at room temperature, cell fixation and permeabilization with BD cytofix/cytoperm kit (BD Biosciences) for 20 min at room temperature, and then intracellular stain for 20 min at room temperature. B cell and monocyte surface staining was performed on freshly-thawed PBMCs with no rest as previously described (Oneko et al., 2021). See KEY RESOURCES TABLE for a complete list of antibodies used. Upon completion of staining, cells were collected on a BD FACSymphony flow cytometer (BD Biosciences, San Jose, CA). Samples were analyzed using FlowJo 10.6.1 (TreeStar). Anomalous “bad” events were separated from “good” events using FlowAI (Monaco et al., 2016). “Good events” were used for all downstream gating. Gating strategies have been previously reported (Oneko et al., 2021).

### Class prediction

The xgboost machine learning algorithm was used to determine which baseline, post-vaccination, or post-vaccination correcting for baseline (delta, Δ) features best predicted parasitemia outcomes at 3 months for individuals who received the 1.8×10^6^ PfSPZ vaccine (n=63 for baseline, n=62 for post-vaccination, and n=61 for Δ). For the baseline analysis, RNA-seq transcriptomic features, 10 flow cytometry features, 15 multiplex plasma cytokine features, anti-CSP IgG antibody reactivity, and 11 in vitro cell stimulation features were used as input. For the post-vaccination analysis the log_2_ fold-change of anti-CSP response, and 10 flow cytometry features, and transcriptomic data from two weeks post-vaccination were used as input. RNA-seq transcriptomic features were collapsed into module expression scores (MES) by determining median expression of all genes within each previously defined low-annotation blood transcription module (346 features) (Li et al., 2014). The following procedures were performed separately for 1) baseline features using MES transcriptomic features only, 2) baseline features using MES transcriptomic features in addition to cell population, CSP-specific IgG, and plasma cytokine data, 3) post-vaccination using MES transcriptomic features only, 4) post-vaccination using MES transcriptomic features in addition to cell population and CSP-specific IgG, 5) delta model using MES transcriptomic features, and 6) delta model using MES transcriptomic features as well as cell population and CSP-specific IgG data. Individuals that were missing more than two-thirds of feature data were removed prior to imputation with the package missMDA (Josse and Husson, 2016). Missing values were imputed by feature for remaining subjects. Two-thirds of the individuals (n=42 for baseline, n=41 for post-vaccination, n=41 for Δ) were randomly selected as the training set and used for feature selection and model training with the remaining one-third held out for independent testing. All feature selection and modeling were performed in R. The caret package was used to perform recursive feature elimination (rfe) on the training set until features were reduced to the top 3 to 5 most predictive features (Kuhn, 2020). Final feature count (3, 4, or 5) for each model was based on performance using a random forest algorithm as part of the rfe command. This included 25 rounds of bootstrapping and was repeated 5 times to account for variability due to random selection with the most commonly repeated features used in the final models. Kappa values were used to evaluate the performance of models based on these selected features. Ideal model parameters were determined using logistic regression over 1000 rounds of cross-validation using the package xgboost (Chen and Guestrin, 2016). Parameters were further tuned manually to maximize predictive accuracy while minimizing overfitting. Seeds were recorded for reproducibility. Final models were then validated using the one-third of samples (n=21 for baseline, n=21 for post-vaccination, and n=20 for Δ) originally held back as test sets. The results of this model prediction were used to generate the confusion matrix and receiver operating characteristics curve using the prOC package, as well as to determine feature importance. Shapley additive explanations (SHAP) dependence plots were generated to determine the contribution of each feature to model prediction output.

### Correlation analysis of transcriptomic and non-transcriptomic data

Correlations were made for transcriptomic data and non-transcriptomic data at baseline, post-vaccination, and ‘delta’ (fold-change between post-vaccination and baseline status). Data was presented as chord diagrams. For transcriptomic data, RNA-seq results were collapsed into MES based on either high-annotation blood transcription modules or modules derived from the Monaco et al. RNA-seq dataset (see above). Non-transcriptomic parameters included FACS data, post-vaccination CSP-specific IgG responses, and time to parasitemia at 6 months post-vaccination. For baseline analysis, we also included cytokine levels as determined by multiplex cytokine assay (see above). Correlation analysis was performed using the ggcorrplot and visualized using the circlize package (Gu et al., 2014) in R. Only positive correlations that fell below FDR values indicated in the figure legends were included in the chord diagrams.

### Sporozoite phagocytosis assay

The sporozoite phagocytosis assay used in this study was modified from a previously described protocol (Steel et al., 2017). Briefly, THP-1 monocytic cells (American Type Culture Collection) were cultured in RPMI medium containing 10% fetal bovine serum (cRPMI) supplemented with either LPS from *Escherichia coli* K12 (2 μg/ml), polyinosinic:polycytidylic acid (poly I:C, 10 μg/ml), flagellin from *Salmonella typhimurium* (1 μg/ml), imiquimod (2 μg/ml), CpG ODN 2006 (5 μM), or β-glucan (100 μg/ml) for 22 to 36 h at 37°C in 5% CO_2_ at a final concentration of 1×10^5^ cells per well in 96-well U bottom plates. Unstimulated cells (medium only) were used as a control. All conditions were done in triplicate wells. Cells were washed with cRPMI and resuspended in fresh medium (15 μl/well). Approximately 1×10^5^ of freshly isolated, motile sporozoites were added to the pre-treated THP-1 cells at a sporozoite to monocyte ratio of 1:3, centrifuged at 500 g for 3 min, and incubated for 1 h at 37°C in 5% CO_2_. Cells were immediately fixed using ice-chilled eBioscience FoxP3/Transcription Factor Staining Buffer Set (ThermoFisher), incubated on ice for 10 min, washed with phosphate-buffered saline with 5 mM EDTA (PBS-EDTA). Cells were centrifuged at 500 g for 5 min. After removing the supernatant, cells were blocked with 2% bovine serum albumin (BSA) in the permeabilization/wash buffer and incubated for 10 min at room temperature with gentle agitation. Cells were washed with PBS-EDTA and stained with 2F6, a neutralizing mouse monoclonal antibody that binds the PyCSP repeat region (Sack et al., 2014), and incubated overnight at 4°C with gentle agitation. Cells were washed twice with PBS-EDTA and incubated with an AlexaFluor 488-conjugated goat anti-mouse IgG secondary antibody (Invitrogen) in a permeabilization/wash buffer containing 2% BSA for 45 min at room temperature. Cells were washed twice, stained with nuclear stain (Hoechst) for 10 min, washed again, resuspended in PBS-EDTA, and acquired on an Imagestream flow cytometer (Luminex).

Analysis of phagocytosis was performed using IDEAS software version 6.0 (Amnis). Briefly, 30,000 cells were acquired per sample. After exclusion of doublets, debris, and dead cells, the gating strategy outlined in Figure S7B was employed. Hoechst^+^ THP-1 cells were determined for the presence of sporozoite internalization. To exclude noise and accurately discriminate between cells with cell-surface bound (external) sporozoites versus phagocytosed (internal) sporozoites within the Hoechst^+^PyCSP^+^ gate (R3), the “Feature Finder Wizard” was used to manually train two populations within R3 by selecting cell images with external (Population 1, n=20) or internal sporozoites (Population 2, n=20) and then randomly selected a feature from the default feature lists (e.g. location, shape, size, texture, signal strength) to find features that best differentiate the assigned populations. A comparison feature describing the difference of intensity measurements between masks or pixels in different images provided the best separation between the assigned populations and was subsequently applied to all samples. Results were reported as internal Hoechst^+^PyCSP^+^/all Hoechst^+^ cells. Counts across all replicates and experiments for each treatment condition (e.g. LPS, poly I:C) were combined into a final analysis comparing proportion of internal Hoechst^+^PyCSP^+^/all Hoechst^+^ cells in treatment versus media only control. Significance and odds ratio was determined by Fisher’s exact test.

### Mouse studies

#### Liver parasite burden determination in mice

Mice that received viable sporozoites were anesthetized ∼42 h post-injection using 3.5% isoflurane, 1.5 L/min O_2_ and were then cervically dislocated. Livers were removed using aseptic technique and placed in 2 mL of RNAlater Solution (Invitrogen, Cat. AM7020). The left median lobe was dissected and weighed prior to bead mill homogenization (5.3 m/s for 45 sec; Bead Ruptor Elite, Omni International; 17 × 2.8mm Ceramic beads, Omni International, Cat. 19-646-3) in 1 mL RLT buffer (RNeasy Plus Mini Kit, Qiagen, Cat. 74134). Liver homogenates were placed on ice prior to proceeding with RNA extraction using the RNeasy Plus Mini Kit (Qiagen) according to the manufacturer’s protocol, optimized for liver tissue. Purity of the RNA samples was assessed via Nanodrop One (ThermoFisher), followed by cDNA synthesis (ProtoScript II Reverse Transcriptase, New England Biolabs, Cat. M0368L; Random Primer Mix, New England Biolabs, Cat. S1330S; dNTPs, Genscript, Cat. C01689; SUPERase-In RNase Inhibitor, Invitrogen, Cat. AM2694) using the manufacturer’s quick protocol for random primer mix. cDNA was amplified using *P. yoelii* 18S primers listed in the KEY RESOURCES TABLE. Real-time PCR was used to quantify relative transcript abundance in the samples using a standard curve for the 18S PCR generated with *P. yoelii* stabilite reference cDNA. Express PrimeTime 5’ 6-FAM(tm)/ZEN(tm)/3’ IB®FQ chemistry was used for the 18S PCR (Integrated DNA Technologies, refer to KEY RESOURCES TABLE) and SYBR chemistry was used for the GAPDH PCR (Luna, New England Biolabs, Cat. M3003S) on an Applied Biosystems QuantStudio 6 Flex Real-Time PCR System (ThermoFisher). Peripheral blood from anesthetized mice was collected 24 h prior to RAS injection, and on days 5, 6, 7, 14, 28, and 55 post-RAS-injection. Red blood cells were lysed and leukocytes were stained with Zombie Aqua (Biolegend, Cat. 423102), washed, resuspended in FACS buffer (PBS with 1% heat inactivated FBS; Atlanta Biologicals, Cat. S11550, 0.02% sodium azide; VWR, Cat. BDH7465-2) containing FC block (anti-CD16/32; clone 2.4G2) for 10 min and stained for 20 min with antibodies listed in the KEY RESOURCES TABLE. Cells were washed, fixed for 20 min (Fixation Buffer; Biolegend, Cat. 420801), and then washed again. Labelled cells were acquired on the BD LSRFortessa X-20 cytometer and data was analyzed using FlowJo software (v.10.7.1) (TreeStar).

## QUANTIFICATION AND STATISTICAL ANALYSIS

Statistical analyses were performed using R software (version 3.5.1) or GraphPad Prism (version 9.1.0). Specific tests for statistical significance are described in figure and table legends. A p value of <0.05 was considered statistically significant unless otherwise indicated.

## Supporting information

Key Resource Table

## Data Availability

DATA AND CODE AVAILABILITY
The RNA-seq data and metadata for the KSPZV1, VRC312, and VRC314 analyses are available on the database of Genotypes and Phenotypes (dbGaP) under the respective accession numbers: phs002196.v1.p1, phs002422.v1.p1, phs002423.v1.p1. Due to lack of consent from the KSPZV1 participants for storage of sequences on a public repository, sequence-level data for this study will be made available upon request. Reproducible code for analysis and visualizations is available at https://github.com/TranLab/kspzv1-systems-analysis.

https://www.kspzv1.malariasystems.org

https://github.com/TranLab/kspzv1-systems-analysis

## DATA VISUALIZATION

Plots were generated using the following R packages: ggplot2, corrplot, ComplexHeatmap, pheatmap, circlize, and ggpubr. Counts were transformed to log_2_(count-per-million) (logCPM) prior to visualization of gene expression data. The web app was developed using the shiny package. A data visualization web app for the transcriptomic data can be accessed at https://www.kspzv1.malariasystems.org.

## DATA AND CODE AVAILABILITY

The RNA-seq data and metadata for the KSPZV1, VRC312, and VRC314 analyses are available on the database of Genotypes and Phenotypes (dbGaP) under the respective accession numbers: phs002196.v1.p1, phs002422.v1.p1, phs002423.v1.p1. Due to lack of consent from the KSPZV1 participants for storage of sequences on a public repository, sequence-level data for this study will be made available upon request. Reproducible code for analysis and visualizations is available at https://github.com/TranLab/kspzv1-systems-analysis.

### Disclaimer

The findings and conclusions in this report are those of the authors and do not necessarily represent the official position of the Centers for Disease Control and Prevention. The content is solely the responsibility of the authors and does not necessarily represent the official views of the National Institutes of Health.

## ACKNOWLEDGEMENTS

We thank the study participants and the VRC312, VRC314, and KSPZV1 study teams. We thank Peter Crompton for critically reading the manuscript. This work was supported by the Doris Duke Charitable Foundation Clinical Scientist Development Award (Grant #2018091 to TMT) and the National Institutes of Health Vaccine Research Center. Grant #2018091 was not used to carry out the mouse experiments in this study. TMT was also supported by supported by grant K08AI125682 (National Institute of Allergy and Infectious Diseases). In vivo mouse experiments were supported by Indiana University Health – Indiana University School of Medicine Strategic Research Initiative (NWS). The Indiana University Melvin and Bren Simon Cancer Center Flow Cytometry Resource Facility is funded in part by NIH, National Cancer Institute grant P30 CA082709, National Institute of Diabetes and Digestive and Kidney Diseases (NIDDK) grant U54 DK106846, and by NIH instrumentation grant 1S10D012270. Sequencing experiments for the KSPZV1 samples were carried out in the Center for Medical Genomics at Indiana University School of Medicine, which is partially supported by the Indiana University Precision Health Initiative.

## AUTHOR CONTRIBUTIONS

T.M.T. and R.A.S. conceived the project. L.S., J.B., M.L., P.A.S., M.D.M., E.G., R.P., B.J.F., and K.B.Y. performed the experiments. M.L., R.P., and N.W.S designed, performed, and analyzed the mouse experiments. J.B., R.P., E.G., P.A.S., D.N.S., and T.M.T. designed, performed, and analyzed the cellular experiments. K.B.Y., K.B., B.J.F., A.Y., and N.H. sequenced and processed the VRC datasets. M.D.M., L.S., A.U., E.S., H.G., X.X., Y.L. sequenced and processed the KSPZV1 datasets. L.S., E.G., and A.L.O. performed validation experiments for the KSPZV1 RNA-seq gene expression. L.C.S., M.O., K.O., S.K., S.L.H., and R.A.S designed and conducted the clinical trial. R.E.W., L.C.S., and T.M.T analyzed the clinical trials data. T.M.T, L.S., X.L., P.H., and A.U. performed the integrated analyses, machine learning, and data visualizations. T.M.T., L.S., J.B., M.L., R.P., P.H., N.W.S., L.C.S., M.O., and R.A.S. co-wrote the manuscript.

## REFERENCES

Abeler-Dorner, L., Laing, A.G., Lorenc, A., Ushakov, D.S., Clare, S., Speak, A.O., Duque-Correa, M.A., White, J.K., Ramirez-Solis, R., Saran, N., et al. (2020). High-throughput phenotyping reveals expansive genetic and structural underpinnings of immune variation. Nat Immunol 21, 86–100.

Beddall, M., Chattopadhyay, P.K., Kao, S.F., Foulds, K., and Roederer, M. (2016). A simple tube adapter to expedite and automate thawing of viably frozen cells. J Immunol Methods 439, 74–78.

Chen, J., Xu, W., Zhou, T., Ding, Y., Duan, J., and Huang, F. (2009). Inhibitory role of toll-like receptors agonists in Plasmodium yoelii liver stage development. Parasite Immunol 31, 466–473.

Chen, T., and Guestrin, C. (2016). XGBoost: A Scalable Tree Boosting System. In Proceedings of the 22nd ACM SIGKDD International Conference on Knowledge Discovery and Data Mining (San Francisco, California, USA: Association for Computing Machinery), pp. 785–794.

Clyde, D.F. (1975). Immunization of man against falciparum and vivax malaria by use of attenuated sporozoites. Am J Trop Med Hyg 24, 397–401.

Csardi, G., and Nepusz, T. (2006). The igraph software package for complex network research.

Datoo, M.S., Natama, M.H., Some, A., Traore, O., Rouamba, T., Bellamy, D., Yameogo, P., Valia, D., Tegneri, M., Ouedraogo, F., et al. (2021). Efficacy of a low-dose candidate malaria vaccine, R21 in adjuvant Matrix-M, with seasonal administration to children in Burkina Faso: a randomised controlled trial. Lancet 397, 1809–1818.

Denzel, A., Maus, U.A., Rodriguez Gomez, M., Moll, C., Niedermeier, M., Winter, C., Maus, R., Hollingshead, S., Briles, D.E., Kunz-Schughart, L.A., et al. (2008). Basophils enhance immunological memory responses. Nat Immunol 9, 733–742.

Dobano, C., Sanz, H., Sorgho, H., Dosoo, D., Mpina, M., Ubillos, I., Aguilar, R., Ford, T., Diez-Padrisa, N., Williams, N.A., et al. (2019). Concentration and avidity of antibodies to different circumsporozoite epitopes correlate with RTS,S/AS01E malaria vaccine efficacy. Nat Commun 10, 2174.

Epstein, J.E., Tewari, K., Lyke, K.E., Sim, B.K., Billingsley, P.F., Laurens, M.B., Gunasekera, A., Chakravarty, S., James, E.R., Sedegah, M., et al. (2011). Live attenuated malaria vaccine designed to protect through hepatic CD8(+) T cell immunity. Science 334, 475–480.

Fourati, S., Cristescu, R., Loboda, A., Talla, A., Filali, A., Railkar, R., Schaeffer, A.K., Favre, D., Gagnon, D., Peretz, Y., et al. (2016). Pre-vaccination inflammation and B-cell signalling predict age-related hyporesponse to hepatitis B vaccination. Nat Commun 7, 10369.

Fumagalli, F., Noack, J., Bergmann, T.J., Cebollero, E., Pisoni, G.B., Fasana, E., Fregno, I., Galli, C., Loi, M., Solda, T., et al. (2016). Corrigendum: Translocon component Sec62 acts in endoplasmic reticulum turnover during stress recovery. Nat Cell Biol 19, 76.

Gaudinski, M.R., Berkowitz, N.M., Idris, A.H., Coates, E.E., Holman, L.A., Mendoza, F., Gordon, I.J., Plummer, S.H., Trofymenko, O., Hu, Z., et al. (2021). A Monoclonal Antibody for Malaria Prevention. N Engl J Med.

Gewirtz, A.T., Navas, T.A., Lyons, S., Godowski, P.J., and Madara, J.L. (2001). Cutting edge: bacterial flagellin activates basolaterally expressed TLR5 to induce epithelial proinflammatory gene expression. J Immunol 167, 1882–1885.

Gordon, S. (2016). Phagocytosis: An Immunobiologic Process. Immunity 44, 463–475.

Gramzinski, R.A., Doolan, D.L., Sedegah, M., Davis, H.L., Krieg, A.M., and Hoffman, S.L. (2001). Interleukin-12-and gamma interferon-dependent protection against malaria conferred by CpG oligodeoxynucleotide in mice. Infect Immun 69, 1643–1649.

Gross, O., Gewies, A., Finger, K., Schafer, M., Sparwasser, T., Peschel, C., Forster, I., and Ruland, J. (2006). Card9 controls a non-TLR signalling pathway for innate anti-fungal immunity. Nature 442, 651–656.

Gu, Z., Gu, L., Eils, R., Schlesner, M., and Brors, B. (2014). circlize Implements and enhances circular visualization in R. Bioinformatics 30, 2811–2812.

Hagan, T., Cortese, M., Rouphael, N., Boudreau, C., Linde, C., Maddur, M.S., Das, J., Wang, H., Guthmiller, J., Zheng, N.Y., et al. (2019). Antibiotics-Driven Gut Microbiome Perturbation Alters Immunity to Vaccines in Humans. Cell 178, 1313–1328 e1313.

Horos, R., Ijspeert, H., Pospisilova, D., Sendtner, R., Andrieu-Soler, C., Taskesen, E., Nieradka, A., Cmejla, R., Sendtner, M., Touw, I.P., et al. (2012). Ribosomal deficiencies in Diamond-Blackfan anemia impair translation of transcripts essential for differentiation of murine and human erythroblasts. Blood 119, 262–272.

Huan, C., Kelly, M.L., Steele, R., Shapira, I., Gottesman, S.R., and Roman, C.A. (2006). Transcription factors TFE3 and TFEB are critical for CD40 ligand expression and thymus-dependent humoral immunity. Nat Immunol 7, 1082–1091.

Ishizuka, A.S., Lyke, K.E., DeZure, A., Berry, A.A., Richie, T.L., Mendoza, F.H., Enama, M.E., Gordon, I.J., Chang, L.J., Sarwar, U.N., et al. (2016). Protection against malaria at 1 year and immune correlates following PfSPZ vaccination. Nat Med 22, 614–623.

Jagannathan, P., Lutwama, F., Boyle, M.J., Nankya, F., Farrington, L.A., McIntyre, T.I., Bowen, K., Naluwu, K., Nalubega, M., Musinguzi, K., et al. (2017). Vdelta2+ T cell response to malaria correlates with protection from infection but is attenuated with repeated exposure. Sci Rep 7, 11487.

Jentho, E., Novakovic, B., Ruiz-Moreno, C., Kourtzelis, I., Martins, R., Chavakis, T., Soares, M.P., Kalafati, L., Guerra, J., Roestel, F., et al. (2019). Heme induces innate immune memory. bioRxiv, 2019.2012.2012.874578.

Johnson, D.E., O’Keefe, R.A., and Grandis, J.R. (2018). Targeting the IL-6/JAK/STAT3 signalling axis in cancer. Nat Rev Clin Oncol 15, 234–248.

Jongo, S.A., Church, L.W.P., Mtoro, A.T., Chakravarty, S., Ruben, A.J., Swanson, P.A., Kassim, K.R., Mpina, M., Tumbo, A.M., Milando, F.A., et al. (2019). Safety and Differential Antibody and T-Cell Responses to the Plasmodium falciparum Sporozoite Malaria Vaccine, PfSPZ Vaccine, by Age in Tanzanian Adults, Adolescents, Children, and Infants. Am J Trop Med Hyg 100, 1433–1444.

Jongo, S.A., Church, L.W.P., Mtoro, A.T., Schindler, T., Chakravarty, S., Ruben, A.J., Swanson, P.A., Kassim, K.R., Mpina, M., Tumbo, A.M., et al. (2020). Increase of Dose Associated With Decrease in Protection Against Controlled Human Malaria Infection by PfSPZ Vaccine in Tanzanian Adults. Clin Infect Dis 71, 2849–2857.

Josse, J., and Husson, F. (2016). missMDA: A Package for Handling Missing Values in Multivariate Data Analysis. 2016 70, 31.

Kalantari, P., DeOliveira, R.B., Chan, J., Corbett, Y., Rathinam, V., Stutz, A., Latz, E., Gazzinelli, R.T., Golenbock, D.T., and Fitzgerald, K.A. (2014). Dual engagement of the NLRP3 and AIM2 inflammasomes by plasmodium-derived hemozoin and DNA during malaria. Cell Rep 6, 196–210.

Kanehisa, M., Furumichi, M., Sato, Y., Ishiguro-Watanabe, M., and Tanabe, M. (2021). KEGG: integrating viruses and cellular organisms. Nucleic Acids Res 49, D545–D551.

Kanehisa, M., Sato, Y., Kawashima, M., Furumichi, M., and Tanabe, M. (2016). KEGG as a reference resource for gene and protein annotation. Nucleic Acids Res 44, D457–462.

Kasturi, S.P., Skountzou, I., Albrecht, R.A., Koutsonanos, D., Hua, T., Nakaya, H.I., Ravindran, R., Stewart, S., Alam, M., Kwissa, M., et al. (2011). Programming the magnitude and persistence of antibody responses with innate immunity. Nature 470, 543–547.

Kawai, T., Takeuchi, O., Fujita, T., Inoue, J., Muhlradt, P.F., Sato, S., Hoshino, K., and Akira, S. (2001). Lipopolysaccharide stimulates the MyD88-independent pathway and results in activation of IFN-regulatory factor 3 and the expression of a subset of lipopolysaccharide-inducible genes. J Immunol 167, 5887–5894.

Kazmin, D., Nakaya, H.I., Lee, E.K., Johnson, M.J., van der Most, R., van den Berg, R.A., Ballou, W.R., Jongert, E., Wille-Reece, U., Ockenhouse, C., et al. (2017). Systems analysis of protective immune responses to RTS,S malaria vaccination in humans. Proc Natl Acad Sci U S A 114, 2425–2430.

Kisalu, N.K., Idris, A.H., Weidle, C., Flores-Garcia, Y., Flynn, B.J., Sack, B.K., Murphy, S., Schon, A., Freire, E., Francica, J.R., et al. (2018). A human monoclonal antibody prevents malaria infection by targeting a new site of vulnerability on the parasite. Nat Med 24, 408–416.

Korotkevich, G., Sukhov, V., Budin, N., Shpak, B., Artyomov, M.N., and Sergushichev, A. (2021). Fast gene set enrichment analysis. bioRxiv, 060012.

Kotliarov, Y., Sparks, R., Martins, A.J., Mule, M.P., Lu, Y., Goswami, M., Kardava, L., Banchereau, R., Pascual, V., Biancotto, A., et al. (2020). Broad immune activation underlies shared set point signatures for vaccine responsiveness in healthy individuals and disease activity in patients with lupus. Nat Med 26, 618–629.

Kramer, A., Green, J., Pollard, J., Jr., and Tugendreich, S. (2014). Causal analysis approaches in Ingenuity Pathway Analysis. Bioinformatics 30, 523–530.

Kroczek, C., Lang, C., Brachs, S., Grohmann, M., Dutting, S., Schweizer, A., Nitschke, L., Feller, S.M., Jack, H.M., and Mielenz, D. (2010). Swiprosin-1/EFhd2 controls B cell receptor signaling through the assembly of the B cell receptor, Syk, and phospholipase C gamma2 in membrane rafts. J Immunol 184, 3665–3676.

Lahaye, X., Gentili, M., Silvin, A., Conrad, C., Picard, L., Jouve, M., Zueva, E., Maurin, M., Nadalin, F., Knott, G.J., et al. (2018). NONO Detects the Nuclear HIV Capsid to Promote cGAS-Mediated Innate Immune Activation. Cell 175, 488–501 e422.

Langfelder, P., and Horvath, S. (2008). WGCNA: an R package for weighted correlation network analysis. BMC Bioinformatics 9, 559.

Langfelder, P., and Horvath, S. (2012). Fast R Functions for Robust Correlations and Hierarchical Clustering. J Stat Softw 46.

Li, S., Rouphael, N., Duraisingham, S., Romero-Steiner, S., Presnell, S., Davis, C., Schmidt, D.S., Johnson, S.E., Milton, A., Rajam, G., et al. (2014). Molecular signatures of antibody responses derived from a systems biology study of five human vaccines. Nat Immunol 15, 195–204.

Li, S., Sullivan, N.L., Rouphael, N., Yu, T., Banton, S., Maddur, M.S., McCausland, M., Chiu, C., Canniff, J., Dubey, S., et al. (2017). Metabolic Phenotypes of Response to Vaccination in Humans. Cell 169, 862–877 e817.

Liberzon, A., Birger, C., Thorvaldsdottir, H., Ghandi, M., Mesirov, J.P., and Tamayo, P. (2015). The Molecular Signatures Database (MSigDB) hallmark gene set collection. Cell Syst 1, 417–425.

Liehl, P., Zuzarte-Luis, V., Chan, J., Zillinger, T., Baptista, F., Carapau, D., Konert, M., Hanson, K.K., Carret, C., Lassnig, C., et al. (2014). Host-cell sensors for Plasmodium activate innate immunity against liver-stage infection. Nat Med 20, 47–53.

Lyke, K.E., Ishizuka, A.S., Berry, A.A., Chakravarty, S., DeZure, A., Enama, M.E., James, E.R., Billingsley, P.F., Gunasekera, A., Manoj, A., et al. (2017). Attenuated PfSPZ Vaccine induces strain-transcending T cells and durable protection against heterologous controlled human malaria infection. Proc Natl Acad Sci U S A 114, 2711–2716.

Mecklenbrauker, I., Saijo, K., Zheng, N.Y., Leitges, M., and Tarakhovsky, A. (2002). Protein kinase Cdelta controls self-antigen-induced B-cell tolerance. Nature 416, 860–865.

Miller, J.L., Sack, B.K., Baldwin, M., Vaughan, A.M., and Kappe, S.H.I. (2014). Interferon-mediated innate immune responses against malaria parasite liver stages. Cell Rep 7, 436–447.

Minkah, N.K., Wilder, B.K., Sheikh, A.A., Martinson, T., Wegmair, L., Vaughan, A.M., and Kappe, S.H.I. (2019). Innate immunity limits protective adaptive immune responses against pre-erythrocytic malaria parasites. Nat Commun 10, 3950.

Miyamoto, A., Nakayama, K., Imaki, H., Hirose, S., Jiang, Y., Abe, M., Tsukiyama, T., Nagahama, H., Ohno, S., Hatakeyama, S., et al. (2002). Increased proliferation of B cells and auto-immunity in mice lacking protein kinase Cdelta. Nature 416, 865–869.

Monaco, G., Chen, H., Poidinger, M., Chen, J., de Magalhaes, J.P., and Larbi, A. (2016). flowAI: automatic and interactive anomaly discerning tools for flow cytometry data. Bioinformatics 32, 2473–2480.

Monaco, G., Lee, B., Xu, W., Mustafah, S., Hwang, Y.Y., Carre, C., Burdin, N., Visan, L., Ceccarelli, M., Poidinger, M., et al. (2019). RNA-Seq Signatures Normalized by mRNA Abundance Allow Absolute Deconvolution of Human Immune Cell Types. Cell Rep 26, 1627–1640 e1627.

Moncunill, G., Scholzen, A., Mpina, M., Nhabomba, A., Hounkpatin, A.B., Osaba, L., Valls, R., Campo, J.J., Sanz, H., Jairoce, C., et al. (2020). Antigen-stimulated PBMC transcriptional protective signatures for malaria immunization. Sci Transl Med 12.

Mordmuller, B., Surat, G., Lagler, H., Chakravarty, S., Ishizuka, A.S., Lalremruata, A., Gmeiner, M., Campo, J.J., Esen, M., Ruben, A.J., et al. (2017). Sterile protection against human malaria by chemoattenuated PfSPZ vaccine. Nature 542, 445–449.

Murphy, S.C., Deye, G.A., Sim, B.K.L., Galbiati, S., Kennedy, J.K., Cohen, K.W., Chakravarty, S., Kc, N., Abebe, Y., James, E.R., et al. (2021). PfSPZ-CVac efficacy against malaria increases from 0% to 75% when administered in the absence of erythrocyte stage parasitemia: A randomized, placebo-controlled trial with controlled human malaria infection. PLoS Pathog 17, e1009594.

Nakaya, H.I., Wrammert, J., Lee, E.K., Racioppi, L., Marie-Kunze, S., Haining, W.N., Means, A.R., Kasturi, S.P., Khan, N., Li, G.M., et al. (2011). Systems biology of vaccination for seasonal influenza in humans. Nat Immunol 12, 786–795.

Nganou-Makamdop, K., and Sauerwein, R.W. (2013). Liver or blood-stage arrest during malaria sporozoite immunization: the later the better? Trends Parasitol 29, 304–310.

Nussenzweig, R.S., Vanderberg, J., Most, H., and Orton, C. (1967). Protective immunity produced by the injection of x-irradiated sporozoites of plasmodium berghei. Nature 216, 160–162.

Oh, J.Z., Ravindran, R., Chassaing, B., Carvalho, F.A., Maddur, M.S., Bower, M., Hakimpour, P., Gill, K.P., Nakaya, H.I., Yarovinsky, F., et al. (2014). TLR5-mediated sensing of gut microbiota is necessary for antibody responses to seasonal influenza vaccination. Immunity 41, 478–492.

Oneko, M., Steinhardt, L.C., Yego, R., Wiegand, R.E., Swanson, P.A., Kc, N., Akach, D., Sang, T., Gutman, J.R., Nzuu, E.L., et al. (2021). Safety, immunogenicity and efficacy of PfSPZ Vaccine against malaria in infants in western Kenya: a double-blind, randomized, placebo-controlled phase 2 trial. Nat Med 27, 1636–1645.

Ono, T., Yamaguchi, Y., Nakashima, H., Nakashima, M., Ishikiriyama, T., Seki, S., and Kinoshita, M. (2021). Lipopolysaccharide preconditioning augments phagocytosis of malaria-parasitized red blood cells by bone marrow-derived macrophages in the liver, thereby increasing the murine survival after Plasmodium yoelii infection. Infect Immun, IAI0024921.

Ozaki, L.S., Gwadz, R.W., and Godson, G.N. (1984). Simple centrifugation method for rapid separation of sporozoites from mosquitoes. J Parasitol 70, 831–833.

Peled, M., Dragovich, M.A., Adam, K., Strazza, M., Tocheva, A.S., Vega, I.E., and Mor, A. (2018). EF Hand Domain Family Member D2 Is Required for T Cell Cytotoxicity. J Immunol 201, 2824–2831.

Rai, D., Pham, N.L., Harty, J.T., and Badovinac, V.P. (2009). Tracking the total CD8 T cell response to infection reveals substantial discordance in magnitude and kinetics between inbred and outbred hosts. J Immunol 183, 7672–7681.

Rinchai, D., Roelands, J., Toufiq, M., Hendrickx, W., Altman, M.C., Bedognetti, D., and Chaussabel, D. (2021). BloodGen3Module: Blood transcriptional module repertoire analysis and visualization using R. Bioinformatics.

Robinson, M.D., McCarthy, D.J., and Smyth, G.K. (2010). edgeR: a Bioconductor package for differential expression analysis of digital gene expression data. Bioinformatics 26, 139–140.

Robinson, M.D., and Oshlack, A. (2010). A scaling normalization method for differential expression analysis of RNA-seq data. Genome Biol 11, R25.

Roland, J., Soulard, V., Sellier, C., Drapier, A.M., Di Santo, J.P., Cazenave, P.A., and Pied, S. (2006). NK cell responses to Plasmodium infection and control of intrahepatic parasite development. J Immunol 177, 1229–1239.

Rts, S.C.T.P. (2015). Efficacy and safety of RTS,S/AS01 malaria vaccine with or without a booster dose in infants and children in Africa: final results of a phase 3, individually randomised, controlled trial. Lancet 386, 31–45.

Schoettler, N., Rodriguez, E., Weidinger, S., and Ober, C. (2019). Advances in asthma and allergic disease genetics: Is bigger always better? J Allergy Clin Immunol 144, 1495–1506.

Seder, R.A., Chang, L.J., Enama, M.E., Zephir, K.L., Sarwar, U.N., Gordon, I.J., Holman, L.A., James, E.R., Billingsley, P.F., Gunasekera, A., et al. (2013). Protection against malaria by intravenous immunization with a nonreplicating sporozoite vaccine. Science 341, 1359–1365.

Sissoko, M.S., Healy, S.A., Katile, A., Omaswa, F., Zaidi, I., Gabriel, E.E., Kamate, B., Samake, Y., Guindo, M.A., Dolo, A., et al. (2017). Safety and efficacy of PfSPZ Vaccine against Plasmodium falciparum via direct venous inoculation in healthy malaria-exposed adults in Mali: a randomised, double-blind phase 1 trial. Lancet Infect Dis 17, 498–509.

Steinhardt, L.C., Richie, T.L., Yego, R., Akach, D., Hamel, M.J., Gutman, J.R., Wiegand, R.E., Nzuu, E.L., Dungani, A., Kc, N., et al. (2020). Safety, Tolerability, and Immunogenicity of Plasmodium falciparum Sporozoite Vaccine Administered by Direct Venous Inoculation to Infants and Young Children: Findings From an Age De-escalation, Dose-Escalation, Double-blind, Randomized Controlled Study in Western Kenya. Clin Infect Dis 71, 1063–1071.

Subramanian, A., Tamayo, P., Mootha, V.K., Mukherjee, S., Ebert, B.L., Gillette, M.A., Paulovich, A., Pomeroy, S.L., Golub, T.R., Lander, E.S., et al. (2005). Gene set enrichment analysis: a knowledge-based approach for interpreting genome-wide expression profiles. Proc Natl Acad Sci U S A 102, 15545–15550.

Sutton, H.J., Aye, R., Idris, A.H., Vistein, R., Nduati, E., Kai, O., Mwacharo, J., Li, X., Gao, X., Andrews, T.D., et al. (2021). Atypical B cells are part of an alternative lineage of B cells that participates in responses to vaccination and infection in humans. Cell Rep 34, 108684.

Takashima, K., Oshiumi, H., Takaki, H., Matsumoto, M., and Seya, T. (2015). RIOK3-mediated phosphorylation of MDA5 interferes with its assembly and attenuates the innate immune response. Cell Rep 11, 192–200.

Tan, J., Cho, H., Pholcharee, T., Pereira, L.S., Doumbo, S., Doumtabe, D., Flynn, B.J., Schon, A., Kanatani, S., Aylor, S.O., et al. (2021). Functional human IgA targets a conserved site on malaria sporozoites. Sci Transl Med 13.

Tan, J., Sack, B.K., Oyen, D., Zenklusen, I., Piccoli, L., Barbieri, S., Foglierini, M., Fregni, C.S., Marcandalli, J., Jongo, S., et al. (2018). A public antibody lineage that potently inhibits malaria infection through dual binding to the circumsporozoite protein. Nat Med 24, 401–407.

Therneau, T.M., and Grambsch, P.M. (2000). Modeling survival data : extending the Cox model (New York: Springer).

Tran, T.M., Li, S., Doumbo, S., Doumtabe, D., Huang, C.Y., Dia, S., Bathily, A., Sangala, J., Kone, Y., Traore, A., et al. (2013). An intensive longitudinal cohort study of Malian children and adults reveals no evidence of acquired immunity to Plasmodium falciparum infection. Clin Infect Dis 57, 40–47.

Tsang, J.S., Schwartzberg, P.L., Kotliarov, Y., Biancotto, A., Xie, Z., Germain, R.N., Wang, E., Olnes, M.J., Narayanan, M., Golding, H., et al. (2014). Global analyses of human immune variation reveal baseline predictors of postvaccination responses. Cell 157, 499–513.

Wang, L.T., Pereira, L.S., Flores-Garcia, Y., O’Connor, J., Flynn, B.J., Schon, A., Hurlburt, N.K., Dillon, M., Yang, A.S.P., Fabra-Garcia, A., et al. (2020). A Potent Anti-Malarial Human Monoclonal Antibody Targets Circumsporozoite Protein Minor Repeats and Neutralizes Sporozoites in the Liver. Immunity 53, 733–744 e738.

Weiss, W.R., Sedegah, M., Beaudoin, R.L., Miller, L.H., and Good, M.F. (1988). CD8+ T cells (cytotoxic/suppressors) are required for protection in mice immunized with malaria sporozoites. Proc Natl Acad Sci U S A 85, 573–576.

White, M.T., Bejon, P., Olotu, A., Griffin, J.T., Riley, E.M., Kester, K.E., Ockenhouse, C.F., and Ghani, A.C. (2013). The relationship between RTS,S vaccine-induced antibodies, CD4(+) T cell responses and protection against Plasmodium falciparum infection. PLoS One 8, e61395.

World Health Organization (2020). World Malaria Report 2020 (World Health Organization).

Yamamoto, M., Sato, S., Hemmi, H., Hoshino, K., Kaisho, T., Sanjo, H., Takeuchi, O., Sugiyama, M., Okabe, M., Takeda, K., et al. (2003). Role of adaptor TRIF in the MyD88-independent toll-like receptor signaling pathway. Science 301, 640–643.

Zaidi, I., Diallo, H., Conteh, S., Robbins, Y., Kolasny, J., Orr-Gonzalez, S., Carter, D., Butler, B., Lambert, L., Brickley, E., et al. (2017). gammadelta T Cells Are Required for the Induction of Sterile Immunity during Irradiated Sporozoite Vaccinations. J Immunol 199, 3781–3788.

Zhen, R., Moo, C., Zhao, Z., Chen, M., Feng, H., Zheng, X., Zhang, L., Shi, J., and Chen, C. (2020). Wdr26 regulates nuclear condensation in developing erythroblasts. Blood 135, 208–219.

Zimmermann, P., and Curtis, N. (2019). Factors That Influence the Immune Response to Vaccination. Clin Microbiol Rev 32.

