## Supplementary Figures for "Innate immune activation restricts priming and protective efficacy of the radiation-attenuated PfSPZ malaria vaccine"

**Affiliations:**
[1] Division of Infectious Diseases, Department of Medicine, Indiana University School of Medicine, Indianapolis, Indiana, 46202, USA
[2] Department of Microbiology and Immunology, Indiana University School of Medicine, Indianapolis, Indiana, 46202, USA
[3] Division of Pediatric Infectious Diseases and Global Health, Department of Pediatrics, Indiana University School of Medicine, Indianapolis, Indiana, 46202, USA
[4] Cellular Immunology Section, Vaccine Research Center, NIAID, NIH, Bethesda, Maryland, 20892, USA
[5] Malaria Branch, Division of Parasitic Diseases and Malaria, Center for Global Health, Centers for Disease Control and Prevention, Atlanta, Georgia, 30333, USA
[6] Broad Institute of MIT Harvard, Cambridge, MA, 02142 USA
[7] Center for Global Infectious Disease Research, Seattle Children's Research Institute, Seattle, WA 98109, USA
[8] Stark Neurosciences Research Institute, Indiana University School of Medicine, Indianapolis, Indiana, 46202, USA
[9] Center for Medical Genomics, Indiana University School of Medicine, Indianapolis, Indiana, 46202, USA
[10] Department of Pediatric Oncology, Dana-Farber Cancer Institute, Boston, MA, USA
[11] Deming Department of Medicine, Tulane University School of Medicine, New Orleans, LA, 70112, USA
[12] Kenya Medical Research Institute, Centre for Global Health Research, Kisumu, Kenya
[13] Sanaria, Rockville, Maryland, 20850, USA
[14]Lead contact

**A**

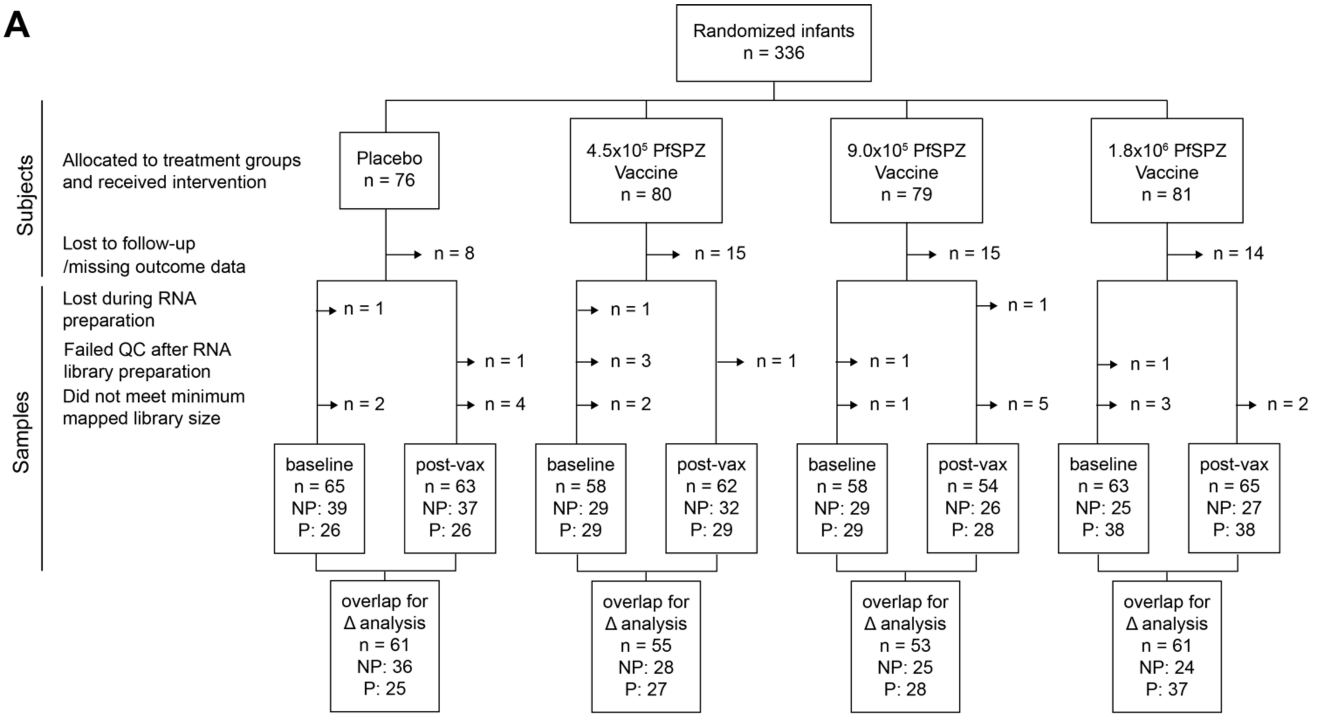


**B**

| | N | not protected (NP) | protected (P) | significance |
|---|---|---|---|---|
| **placebo** | | (N=40) | (N=26) | |
| Site : Wagai | 66 | 17 (42.50%) | 20 (76.92%) | P=0.012[1] |
| Sex : M | 66 | 22 (55.00%) | 13 (50.00%) | P=0.884[1] |
| Age | 66 | 6.42 **9.00** 10.00 | 6.92 **8.50** 11.00 | P=0.806[2] |
| Hemoglobin | 66 | 9.30 **10.00** 10.80 | 9.88 **10.65** 11.32 | P=0.066[2] |
| Platelets | 66 | 278.08 **394.50** 461.75 | 333.17 **412.50** 516.50 | P=0.256[2] |
| Weighted_z | 66 | -0.74 **-0.18** 0.32 | -0.57 **0.29** 0.93 | P=0.126[2] |

| | N | not protected (NP) | protected (P) | significance |
|---|---|---|---|---|
| **4.5x10⁵ PfSPZ** | | (N=30) | (N=29) | |
| Site : Wagai | 59 | 12 (40.00%) | 13 (44.83%) | P=0.911[1] |
| Sex : M | 59 | 18 (60.00%) | 15 (51.72%) | P=0.706[1] |
| Age | 59 | 7.00 **8.00** 11.00 | 6.00 **8.00** 11.00 | P=0.921[2] |
| Hemoglobin | 59 | 9.97 **10.85** 11.40 | 10.30 **10.80** 11.33 | P=0.867[2] |
| Platelets | 59 | 297.08 **376.50** 433.00 | 347.00 **443.00** 557.33 | P=0.058[2] |
| Weighted_z | 59 | -1.14 **-0.24** 0.46 | -1.14 **-0.06** 0.57 | P=0.891[2] |

| | N | not protected (NP) | protected (P) | significance |
|---|---|---|---|---|
| **9.0x10⁵ PfSPZ** | | (N=33) | (N=31) | |
| Site : Wagai | 64 | 17 (51.52%) | 12 (38.71%) | P=0.437[1] |
| Sex : M | 64 | 14 (42.42%) | 14 (45.16%) | P=1.000[1] |
| Age | 64 | 7.00 **8.00** 10.33 | 6.00 **7.00** 9.00 | P=0.166[2] |
| Hemoglobin | 64 | 8.97 **9.70** 10.87 | 10.12 **10.60** 11.27 | P=0.050[2] |
| Platelets | 64 | 260.33 **373.00** 486.67 | 290.00 **444.00** 556.83 | P=0.097[2] |
| Weighted_z | 64 | -1.07 **-0.31** 0.39 | -0.99 **0.05** 0.31 | P=0.904[2] |

**C**

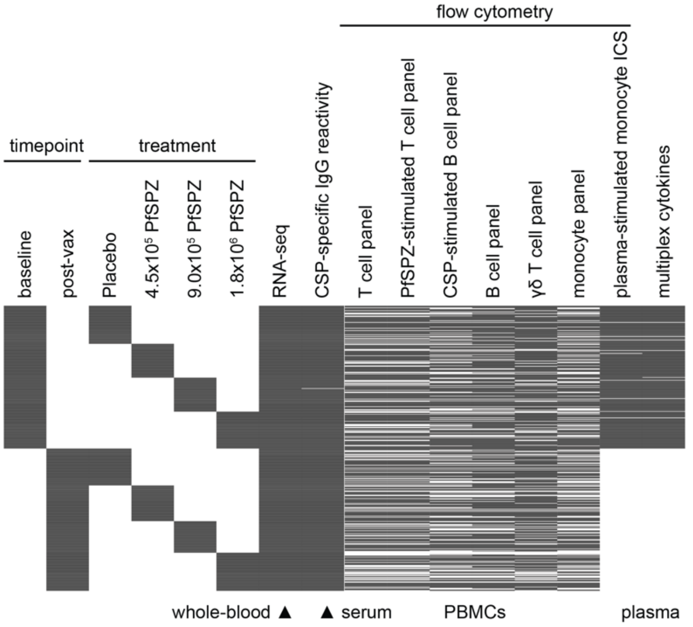


| | N | not protected (NP) | protected (P) | significance |
|---|---|---|---|---|
| **1.8x10⁶ PfSPZ** | | (N=28) | (N=39) | |
| Site : Wagai | 67 | 12 (42.86%) | 21 (53.85%) | P=0.522[1] |
| Sex : M | 67 | 18 (64.29%) | 27 (69.23%) | P=0.872[1] |
| Age | 67 | 6.00 **8.00** 9.58 | 6.33 **9.00** 10.00 | P=0.239[2] |
| Hemoglobin | 67 | 9.60 **10.30** 11.22 | 9.22 **10.40** 11.17 | P=0.819[2] |
| Platelets | 67 | 342.33 **440.50** 555.17 | 322.83 **432.00** 557.33 | P=0.794[2] |
| Weighted_z | 67 | -0.75 **-0.06** 0.57 | -0.79 **-0.16** 0.50 | P=0.576[2] |

**Figure S1. Participants and samples used in study.** Related to Figure 1.
(A) Flow diagram of subjects and samples used for transcriptomic analyses.
(B) Characteristics study participants by outcome and treatment. [1]Two-proportions Z-test and [2]Wilcox test used to determine statistical significance for categorical and continuous variables, respectively.
(C) Samples used for each assay by timepoint and treatment.

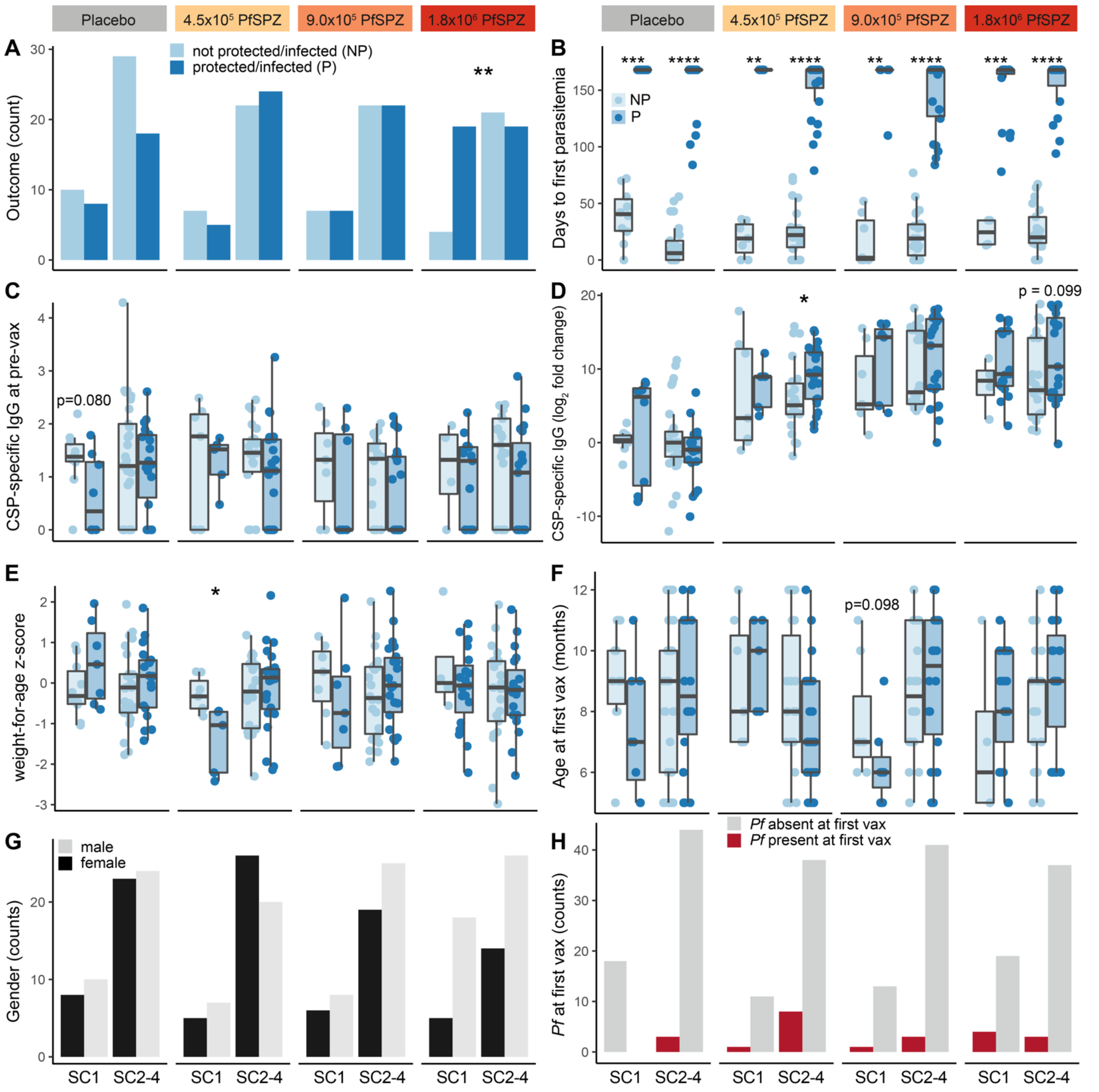


**Figure S2. Comparisons within pre-immunization sample cluster 1 (SC1) and SC2-4 by treatment group.** Related to Figure 1.

(A) Distribution of outcomes between clusters.
(B) Comparison of days to first parasitemia up to 6 months of follow up between outcomes.
(C) Comparison of CSP-specific IgG at pre-immunization baseline (pre-vax) between outcomes.
(D) Comparison of $\log_2$ fold-change in CSP-specific IgG at 2 weeks post-immunization between outcomes.
(E) Comparison of weight-for-age z-score between outcomes.
(F) Comparison of age at first vax between outcomes.
(G) Distribution of gender between clusters.
(H) Distribution of *Pf* parasitemia at first vaccination (vax) between clusters.

Outcomes are not protected/infected (NP; dark blue) or protected/not infected (P; light blue) during 3 months of surveillance post-immunization (see Figure 1A). Sample clusters were based on unsupervised hierarchical clustering of whole-blood transcriptomic profiles at pre-immunization baseline (see Figure 1C). Wilcoxon and Fisher's exact tests used to determine statistical significance for continuous and categorical variables, respectively. Statistical significance indicated by *p<0.05, **p<0.01, ***p<0.001, ****p<0.0001. Non-significant p values shown if 0.05≤p<0.10.

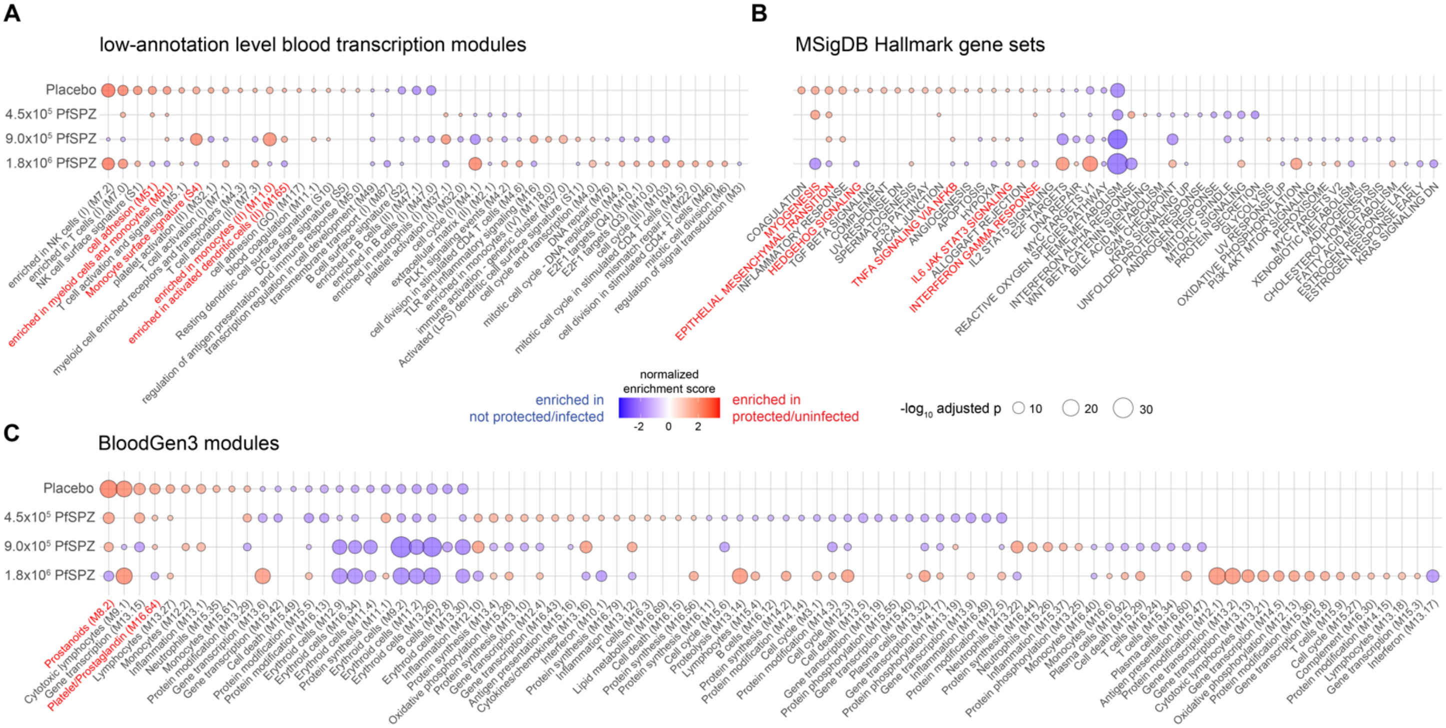


**Figure S3. Gene set enrichment analysis of pre-immunization whole-blood transcriptomes between infants who were protected (uninfected) through 3 months of follow up versus those who were not protected (infected).** Related to Figure 2.

(A) Using low-annotation level blood transcription modules

(B) Using MSigDB Hallmark gene sets

(C) Using BloodGen3 modules

Modules are sorted from left to right by decreasing normalized enrichment score and treatment. Modules enriched in “protected” for Placebo but enriched in “not protected” for 1.8x10$^6$ PfSPZ are highlighted in red text. Only modules with a minimum gene set size of 20 and a Benjamini-Hochberg adjusted p <0.20 are shown.

**A**

| Study | Group | Regimen | Not Protected | Protected |
|---|---|---|---|---|
| VRC 312 | 3a | 30,000 PfSPZ x 6 | 3 | 0 |
| | 3b | 30,000 PfSPZ x 4 | 8 | 1 |
| | 4a | 135,000 PfSPZ x 5 | 0 | 2 |
| | 4b | 135,000 PfSPZ x 5 | 0 | 4 |
| | 4c | 135,000 PfSPZ x 4 | 3 | 6 |
| | All | | 14 | 13 |
| VRC 314 | 1 | 270,000 PfSPZ x 3 | 1 | 3 |
| | 4 | 270,000 PfSPZ x 4 | 0 | 5 |
| | 5 | 270,000 PfSPZ x 4 | 5 | 6 |
| | 6 | 900,000 PfSPZ x 3 | 5 | 9 |
| | All | | 11 | 23 |

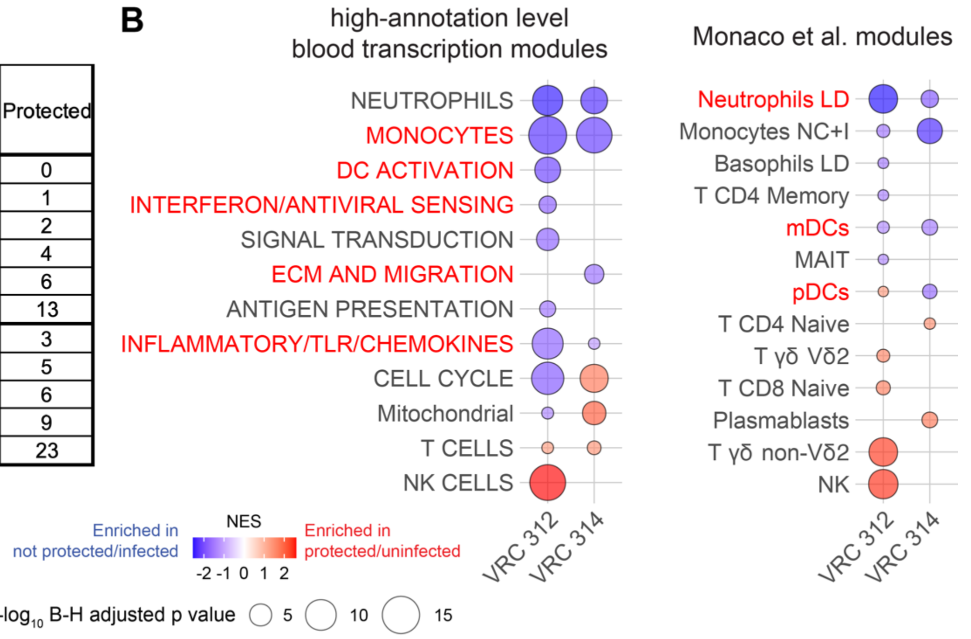


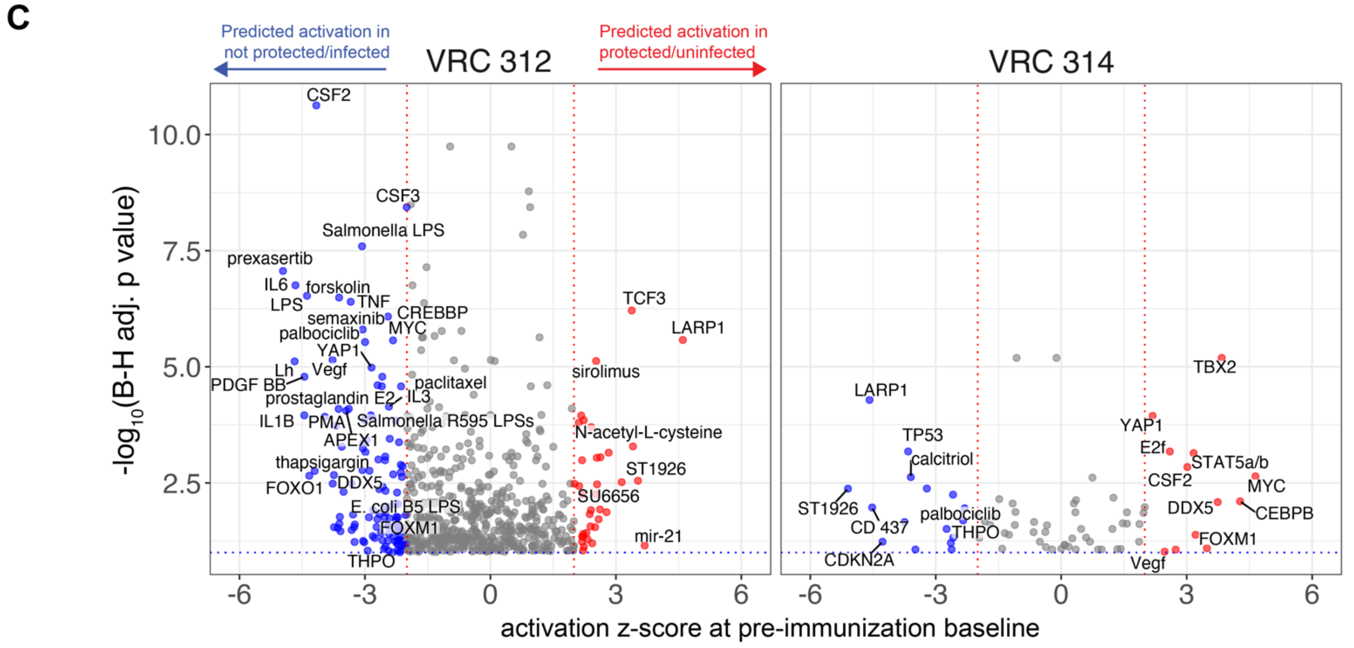


**Figure S4. Differential expression analysis of whole-blood transcriptomes at pre-immunization baseline for VRC 312 and VRC 314 PfSPZ Vaccine trials.** Related to Figure 2.

(A) Numbers of protected and not protected subjects by dose regimen and trial. Only groups in which a consistent PfSPZ dose was delivered by DVI was included for differential gene expression (DGE) analysis.

(B) Gene set enrichment analysis of baseline transcriptomes between protected and not protected subjects

(C) Upstream regulator analysis of baseline transcriptomes between protected and not protected subjects

DGE analyses between outcome groups included adjustments for sequencing batch and dose regimen. B-H = Benjamini-Hochberg; NES = normalized enrichment score. Only modules with a minimum gene set size of 20 and a B-H adjusted p <0.20 are shown.

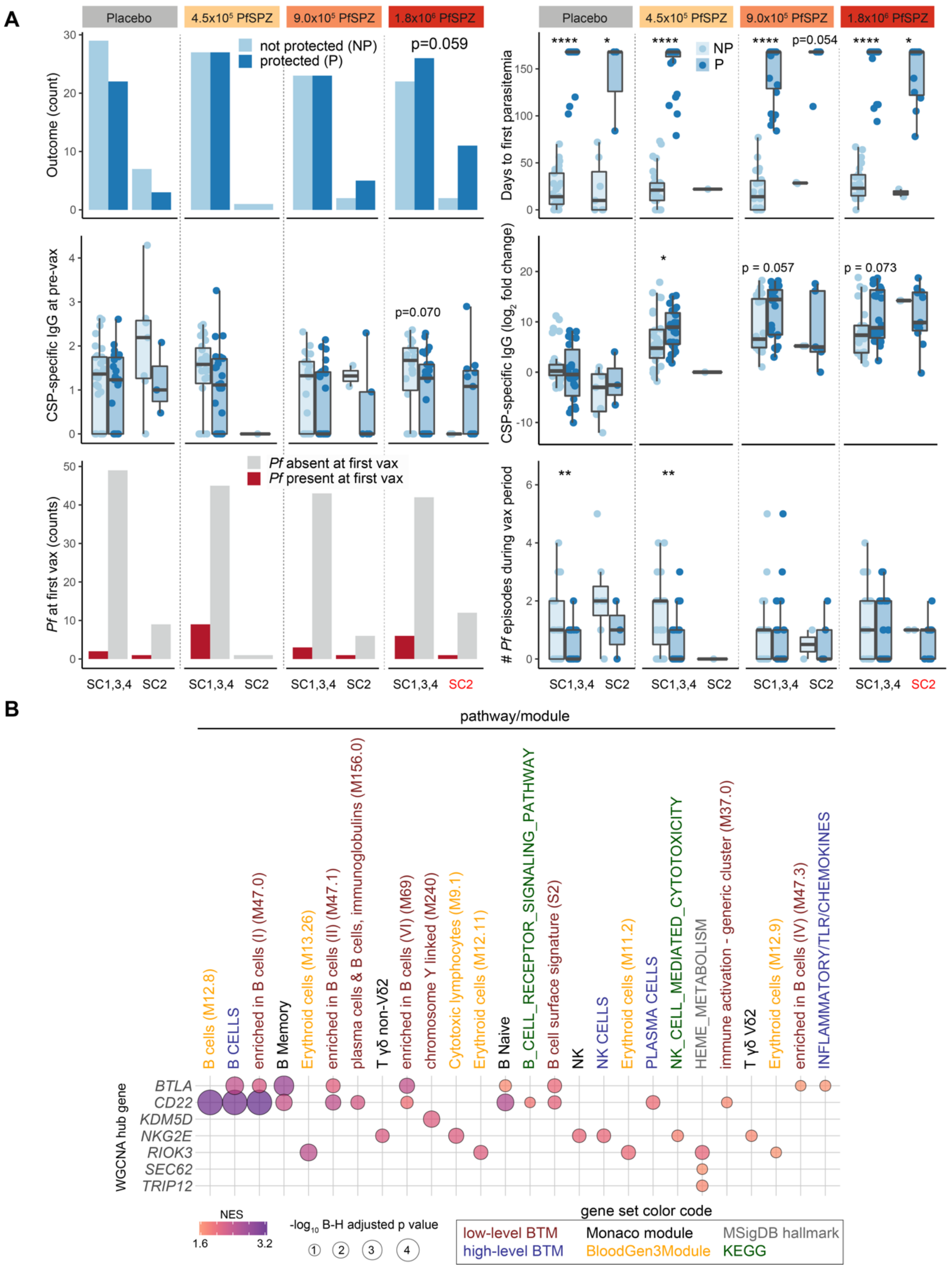


**Figure S5. Cluster analyses of changes in post-immunization whole-blood transcriptomes over pre-immunization baseline.** Related to Figure 4.

(A) Comparisons of indicated variables within delta sample cluster 2 (SC2) and SC1,3,4 by treatment group. Outcomes are not protected/infected (NP; dark blue) or protected/not infected (P; light blue) during 3 months of surveillance post-immunization (see Figure 4B). Sample clusters were based on unsupervised hierarchical clustering of changes in whole-blood transcriptomes over the vaccination period (post-immunization/pre-immunization baseline = Δ). Wilcoxon test and Fisher's exact used to determine statistical significance for continuous and categorical variables, respectively. Significance indicated by *p<0.05, **p<0.01, ***p<0.001, ****p<0.0001. Non-significant p values shown if p<0.10.

(B) Gene set enrichment analysis of genes ranked by membership within indicated modules which were determined by weight gene correlation network analysis (WGCNA) of whole-blood Δ transcriptomes. Related to Figures 4E-F.

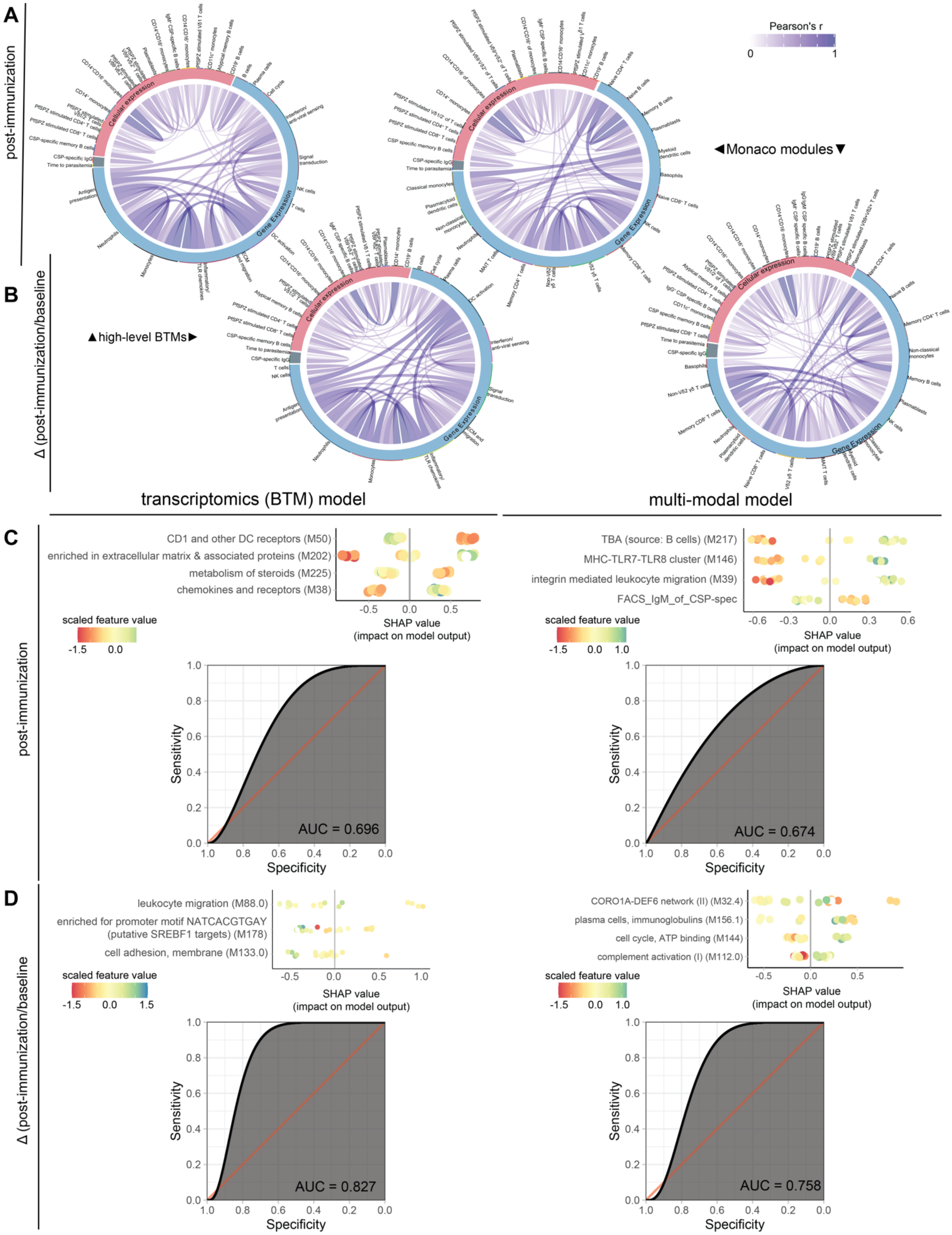


**Figure S6. Integrated correlative and machine learning analyses.** Related to Figure 5.
Chord diagrams showing pairwise correlations between (A) post-immunization or (B) delta (post-immunization/baseline) features (gene expression collapsed into either high-annotation level blood transcription or Monaco modules and flow cytometric features [“cellular expression”]) with the post-vaccination outcomes of plasma CSP-specific IgG and time-to-parasitemia at 6 months for all infants with available data. Blue lines indicate significantly positive correlations (FDR<0.20). SHapley Additive exPlanations (SHAP) plots and ROC curves for indicated models using (C) post-immunization or (D) delta (post-immunization/baseline) features. For SHAP plots, each point is a single subject in the test set. AUC = area under the curve.

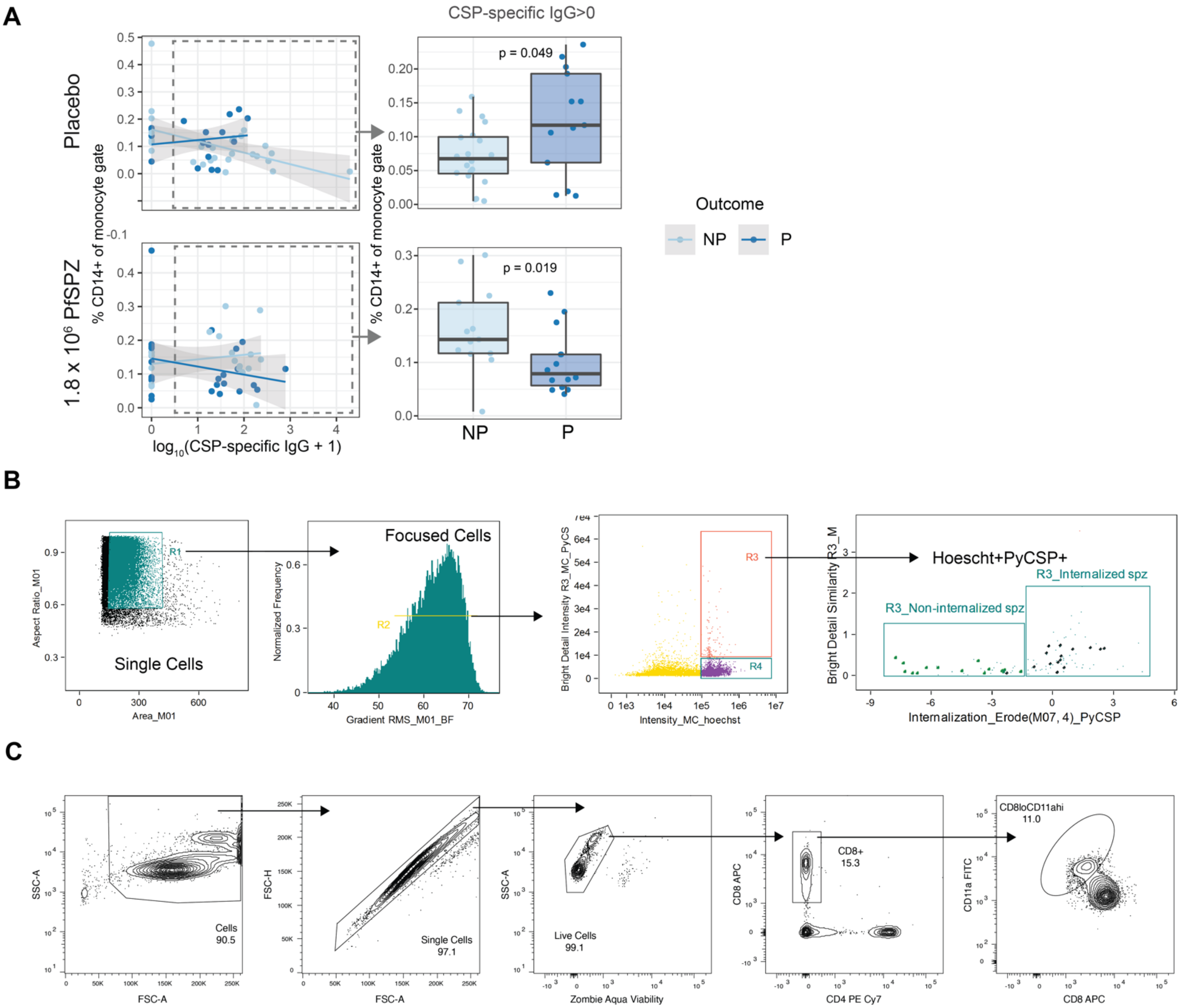


**Figure S7. Relationship between monocytes and CSP-specific IgG and gating strategies for detection of phagocytosed sporozoites and RAS-induced CD8 T cells (CD8loCD11ahi).** Related to Figures 6 and 7.

(A) % CD14+ peripheral monocytes by outcome and treatment for infants with baseline CSP-specific IgG>0 for infants who were not protected (NP) or protected (P) post-vaccination.

(B) Gating strategy for identifying THP-1 cells with internalized *P. yoelii* sporozoites.

(C) Representative flow cytometry plots identifying RAS-induced CD8 T cell (CD8loCD11ahi). Value within plot is the percent of all circulating CD8 T cells that are CD8loCD11ahi.
