## Supplementary material for "Innate immune activation restricts priming and protective efficacy of the radiation-attenuated PfSPZ malaria vaccine": Table S6

| Time Point | Model Type | Features in Model | Number of Runs | Best Iteration | Predictive Accuracy | RMSE | AUC |
| --- | --- | --- | --- | --- | --- | --- | --- |
| Pre-Immunization | Transcriptomic | 3 | 441 | 43 | 71.40% | 28.60% | 76.60% |
| Post-Immunization | Transcriptomic | 4 | 541 | 144 | 72.70% | 27.70% | 69.60% |
| Delta | Transcriptomic | 3 | 401 | 5 | 71.40% | 28.60% | 82.70% |
| Pre-Immunization | Multi-modal | 4 | 671 | 274 | 76.10% | 23.80% | 86.20% |
| Post-Immunization | Multi-modal | 4 | 481 | 81 | 75% | 25% | 67.40% |
| Delta | Multi-modal | 4 | 411 | 11 | 70% | 30% | 75.80% |
